# Characterize Disease Progression Subphenotypes in Real World Populations with Overweight and Obesity using a Graph-based Neural Network Framework

**DOI:** 10.1101/2025.11.10.25339913

**Authors:** Yao An Lee, Yu Huang, Hao Dai, Tarik K. Yuce, Viral Shah, Jiang Bian, Jingchuan Guo

## Abstract

**Background:** Obesity is a chronic, heterogeneous condition, with risks, trajectories, and treatment responses that vary widely among individuals. However, research characterizing the heterogeneity of long-term obesity progression—and its impact on the development of obesity-associated outcomes and treatment responses—is scarce.

**Objectives:** We aimed to identify progression subphenotypes in a real-world population with overweight or obesity over a period of up to 10 years using electronic health records (EHRs) and evaluate the heterogeneity of treatment effects (HTE) of glucagon-like peptide-1 receptor agonists (GLP-1RAs) across these subphenotypes for major obesity-related disease outcomes.

**Methods:** We conducted a retrospective cohort study of adults from OneFlorida+ who were eligible for anti-obesity medication, defined by the presence of a documented diagnosis of obesity (BMI ≥ 30 kg/m²) or a BMI in the range of 27.0–29.9 kg/m² accompanied by at least one weight-related comorbidity. We developed an outcome-oriented graph neural network (GNN)-based model to identify progression subphenotypes of obesity. Within each subphenotype, we emulated a target trial of GLP-1RA users vs non-users, using propensity score matching and Cox proportional hazards models to evaluate obesity-related disease outcomes, including metabolic dysfunction-associated steatotic liver disease (MASLD), major atherosclerotic cardiovascular disease (ASCVD), stroke, heart failure (HF), and chronic kidney disease (CKD).

**Results:** Among 237,103 adults with overweight or obesity, 58.1% were female, with a mean age of 51.8 years. The study population included 48.4% non-Hispanic White (NHW), 27.9% non-Hispanic Black (NHB), and 13.0% Hispanic individuals. Our GNN model identified three distinct progression subphenotypes: a progressive group (43.6%; “*substantial-increase*” BMI trajectory), an intermediate group (33.4%; “*moderate-increase*”), and a stable group (64.4%; “*steady*”). The mean follow-up durations were comparable across the subphenotypes (4.9-5.0 years). The progressive group exhibited greater baseline multimorbidity, higher opioid exposure, and an initially favorable neighborhood socioeconomic status that declined rapidly over time. The subphenotypes differed significantly in their risks of developing obesity-associated comorbidities and in all-cause mortality during follow-up (p < 0.001). GLP-1RA use was associated with a lower HF risk in two subphenotypes (HR (95% CI): 0.75 (0.61-0.91) and 0.74 (0.61-0.91), respectively) and with a possible increase in CKD risk in the progressive group (1.20 (1.02-1.41)).

**Conclusions:** Obesity progression subphenotypes derived from routine EHRs reveal markedly heterogeneity in trajectories, clinical risks, and treatment responses. Distinct BMI trajectory groups highlight the potential of data-driven, phenotype-guided care and expose meaningful variation in GLP-1RA treatment effects. These findings underscore the potential of longitudinal EHR analysis to advance precision obesity medicine and warrant prospective validation of HTE.

## Introduction

Obesity affects over 1 billion people worldwide,^1^ and its rising prevalence makes it a major risk factor for numerous chronic diseases, including type 2 diabetes mellitus (T2D), cardiovascular disease, metabolic dysfunction-associated steatotic liver disease (MASLD), and cancer. Obesity is a chronic, relapsing condition with a complex, longitudinal disease course influenced by multiple comorbidities.^2,3^ Despite this complexity, current therapeutic decisions primarily rely on body mass index (BMI) for all affected individuals, overlooking crucial variations in comorbidity risk profiles and treatment responses. This approach extends to regulatory guidelines, in which authorities (e.g., the U.S. Food and Drug Administration [FDA]) focus on weight reduction as the primary endpoint for clinical trials rather than addressing underlying metabolic dysfunction or individual patient characteristics.^4^

Glucagon-like peptide-1 receptor agonists (GLP-1RAs) are a class of medications used for T2D treatment and obesity management that act through appetite suppression, delayed gastric emptying, and enhanced thermogenesis.^5,6^ However, substantial heterogeneity in treatment effects exists: younger patients, women, and those with higher baseline BMI achieve significantly greater weight loss, whereas patients with T2D demonstrate less weight reduction than those without T2D.^7–10^ Genetic and pharmacogenomic studies provide important mechanistic insights into this heterogeneity,^11^ and these findings underscore the need to account for obesity heterogeneity in clinical decision-making.

Therefore, understanding the underlying obesity course progression trajectories enables clinicians to stratify patients by complication risk and supporting customized medical interventions for obesity management. Moreover, knowing a patient’s specific subphenotype provides more accurate prognostic information than simple BMI cutoffs. Recent research has demonstrated this potential; for example, in a clinical trial, Acosta et al. identified subphenotypes based on gastrointestinal and psychological traits that responded differently to weight-loss medications.^12^ Chami et al.’s gene-discovery analysis identified eight genetic subphenotypes of obesity with distinct pathways, cardiometabolic risk profiles and serum-protein features, confirming obesity’s substantial heterogeneity. However, most of these studies are cross-sectional rather than longitudinal, and the data underlying these findings are not clinically accessible, limiting their application in real-world scenarios and restricting their use in routine care, particularly in less well-equipped settings.

The proliferation of accessible real-world data (RWD), such as electronic health records (EHRs) and administrative claims data, coupled with advancements in artificial intelligence (AI) and machine learning (ML) techniques, has opened new avenues for investigating obesity heterogeneity in a data-driven manner. Beuken et al. applied probabilistic distance clustering to cross-sectional data from individuals with abdominal obesity, identifying three distinct clusters encompassing demographics, lifestyle, biomedical aspects, advanced phenotyping, and social factors.^13^ Anwar et al. used ML-based integrative unsupervised clustering to identify proteomics- and metabolomics-defined subpopulations of individuals living with obesity.^14^ However, obesity is a progressive condition rather than a fixed state. The body changes over time in ways that reinforce weight gain cycles, making management progressively more challenging.^15,16^ Existing methods have struggled to capture these progression characteristics. Therefore, modeling the underlying relationships within patients’ historical health status and introducing outcome-oriented learning approaches are essential for identifying subgroups with homogeneous progression patterns (i.e., comorbidities, medication use, and clinical biomarkers) that are strongly associated with future clinical outcomes.

We developed a graph neural network (GNN)-based framework that leverages a graph structure to model longitudinal EHRs. This design connects patients’ historical visits that share similar clinical characteristics and enables GNNs to effectively learn patient progression patterns as embeddings. By incorporating an outcome-driven strategy, we ensured that extracted embeddings remain clinically relevant to obesity progression. Additionally, this framework utilizes time-series clustering to analyze sequential embeddings learned from GNNs, identifying distinct obesity-progression subphenotypes. We validated our approach in a large-scale EHR cohort from the OneFlorida+ Clinical Research Consortium, identifying three progression subphenotypes. These three subphenotypes show distinct patterns in comorbidities, drug use, and clinical biomarkers. Following a target trial emulation framework, we evaluated whether the identified subphenotypes exhibited heterogeneous treatment responses by conducting analyses of GLP-1RA therapy across each subphenotype. The results demonstrated differential effectiveness of GLP-1RAs for multiple obesity-related outcomes (e.g., cardiovascular, liver, and kidney diseases), highlighting distinct treatment responsiveness among subgroups.

## Methods

### Data source, study design and study population

In this study, we used RWD from OneFlorida+, a clinical research network contributing to the national Patient-Centered Clinical Research Network (PCORnet), which collaborates with a group of 14 health organizations. OneFlorida+ contains robust, longitudinal, patient-level EHRs of 16.8 million patients from Florida, 2.1 million from Georgia, and 1.1 million from Alabama.

This study employed a retrospective cohort study design to characterize trajectories of obesity progression and to evaluate the association of glucagon-like peptide-1 receptor agonists (GLP-1RAs) with subsequent risk of MASLD, major atherosclerotic cardiovascular disease (ASCVD), stroke, heart failure (HF), and chronic kidney disease (CKD) outcomes in real-world patients. **Figure 1** shows an overview of the study cohort construction process. The study population included patients who were (1) eligible for anti-obesity medications (AOMs), defined as having either a documented diagnosis of obesity, a BMI ≥ 30.0 kg/m², or a BMI of 27.0–29.9 kg/m² with at least one obesity-related comorbidity,^17^ and (2) aged 18 years or older at the first recorded obesity diagnosis. Patients were excluded if they had (1) pregnancy, (2) an active malignant neoplasm, or (3) bariatric surgery at any time during the study period (e**Table 1**). The index date for obesity progression was defined as the date of first obesity diagnosis.

**Figure 1.**
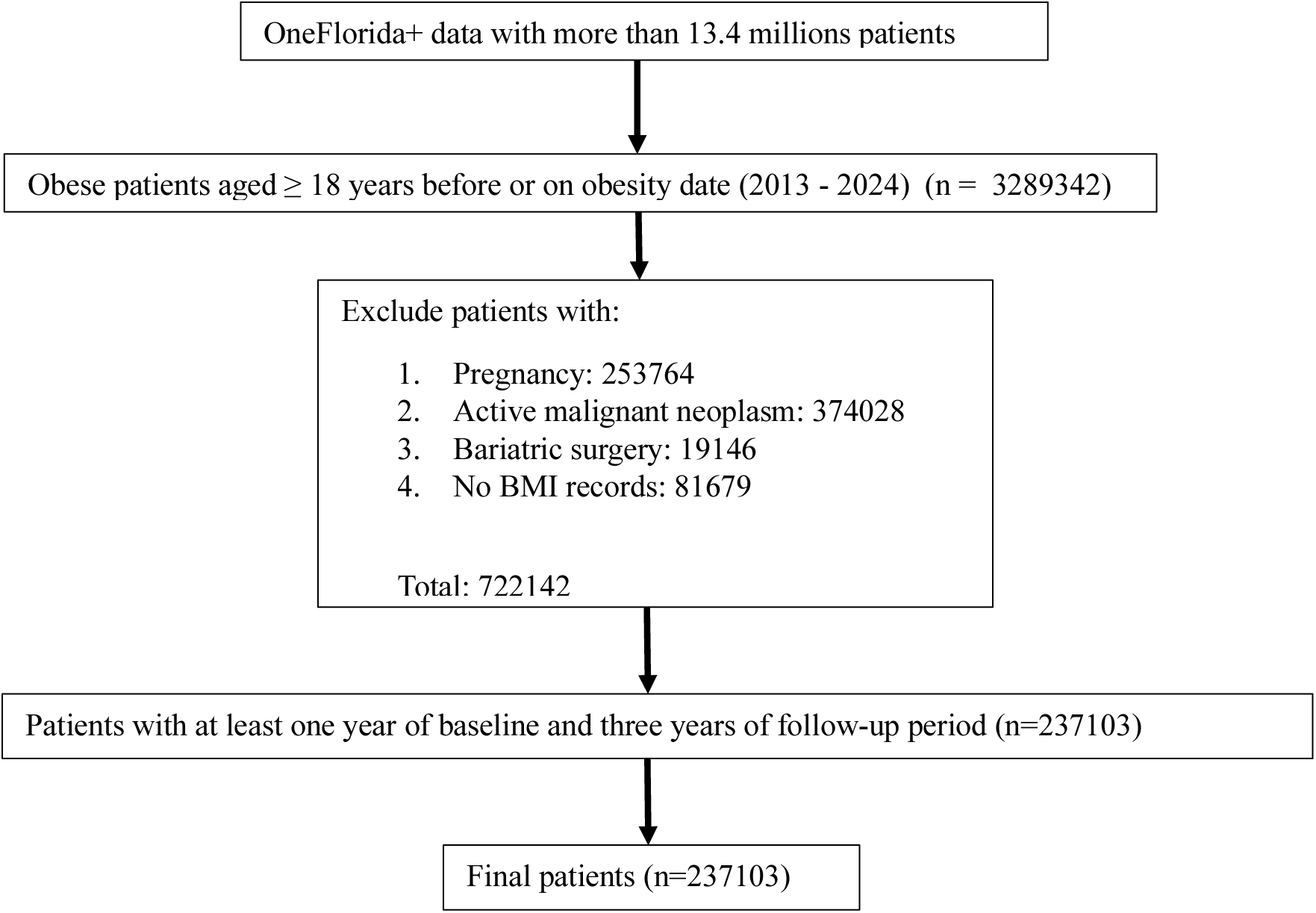
Overview of the study cohort extracted from the OneFlorida+ network.

The index date for GLP-1RA users was defined as the date of the first GLP-1RA prescription. To ensure sufficient observation time for model training, we further restricted the cohort to patients who had at least one year of data before the index date and at least 3 years of follow-up period. These criteria enabled delineation of distinct progression subphenotypes, including a subgroup characterized by severe progression. Within each subphenotype, we then identified GLP-1RA new users as the treatment arm, and non-users as the comparator arm—patients who did not initiate GLP-1RA therapy during the study period. Following a target trial emulation framework, we compared major clinical outcomes between GLP-1RA users and non-users.

### Study outcomes

For the obesity progression subphenotype model, the outcome was defined as the annual change in BMI, calculated as the maximum BMI recorded in the subsequent year minus the minimum BMI of the current year, and categorized into four groups: ≤–1, –1 to 1, 1 to 5, and ≥5 kg/m².

For GLP-1RA target-trial emulation within each subphenotype, the outcomes included MASLD, ASCVD, stroke, HF, and CKD, identified using ICD-9-CM and ICD-10-CM diagnosis codes based on previously validated computable subphenotype algorithms (e**Table 1**).

### Covariates

Baseline covariates encompassed a range of demographic, clinical, pharmacological, and social determinants of health (SDOH) characteristics. Demographic variables included age, sex, race/ethnicity, smoking status, and insurance type. Clinical covariates comprised comorbid conditions (e.g., cardiovascular disease, biliary disorders, and neuropathy), clinical observations (e.g., BMI, blood pressure), laboratory values (e.g., hemoglobin A1c [HbA1c], estimated glomerular filtration rate [eGFR], and lipid profile), and medications (e.g., aspirin, opioids, and statins). SDOH included income level, education level, unemployment rate, median house income, and community food access level (**eTable 2**).

For the GLP-1RA target trial emulation within each subphenotype, the exposure was initiation of GLP-1RA, including semaglutide, tirzepatide, liraglutide, lixisenatide, albiglutide, exenatide, and dulaglutide, ascertained using RxNorm identifiers and National Drug Codes (e**Table 3**). The comparator group comprised patients without evidence of GLP-1RA use during the study period.

### Disease progression subphenotyping framework

Figure 2a illustrates the proposed framework, which consists of four components: (1) Outcome-oriented representation learning using GNNs, (2) Identification of disease progression subphenotypes via time-series clustering, (3) Assessment of subphenotype interpretability by predictive modeling, and (4) Heterogeneity of treatment effects of GLP-1RA across subphenotypes.

**Figure 2.**
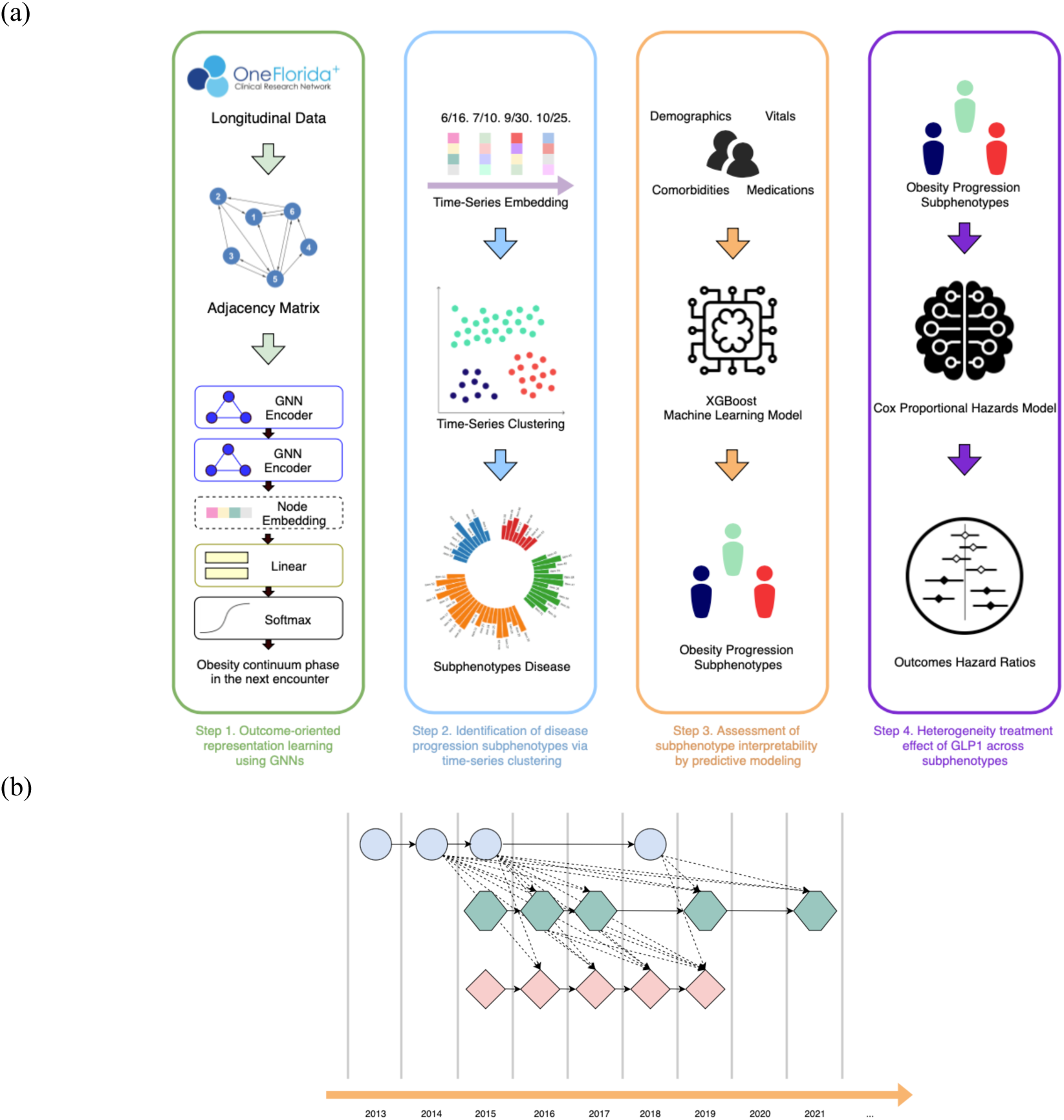
(a) Study pipeline; (b) an example of the proposed Disease Progression Graph (DPG). Each node represents an encounter; the shape and color of a node differentiate the patient, and the dotted lines represent edges that link nodes of different patients.

### Step 1: Outcome-oriented representation learning using GNNs

The longitudinal EHRs of the 𝑛-th patient can be represented as 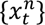, where 𝑡 ∈ {1,2, …, T}, and 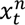 contains multi-source information (e.g., demographics, diagnoses, and treatments) documented at each visit (encounter). For each encounter, we first discretized age into young (18–40), middle (40–65), and old (≥65) and utilized one-hot encoding to encode variables such as age, sex, and race/ethnicity.^18^ We then identified disease diagnoses and medication use at each encounter directly from diagnosis and drug codes (e.g., ICD-9-CM/ICD-10-CM and NDC/RxNorm). We discretized BMI into six categories: normal (<25.0 kg/m²), overweight (25.0–29.9 kg/m²), class 1 obesity (30.0–34.9 kg/m²), class 2 obesity (35.0–39.9 kg/m²), class 3 (morbid) obesity (40.0–49.9 kg/m²), and class 3 (super) obesity (≥50.0 kg/m²),^19^ blood pressure into five classes (normal, elevated, hypertension stage 1, hypertension stage 2, and hypertensive crisis),^20^ and smoking status to current, former, non-smoker, and others. Finally, we formed a binary vector by concatenating the encoded age, gender, and race from the encounters with diagnoses and medications in a one-year period up to time point 𝑡 for each patient (Figure 3a), resulting in an enhanced encounter representation 𝑥-”.

**Figure 3.**
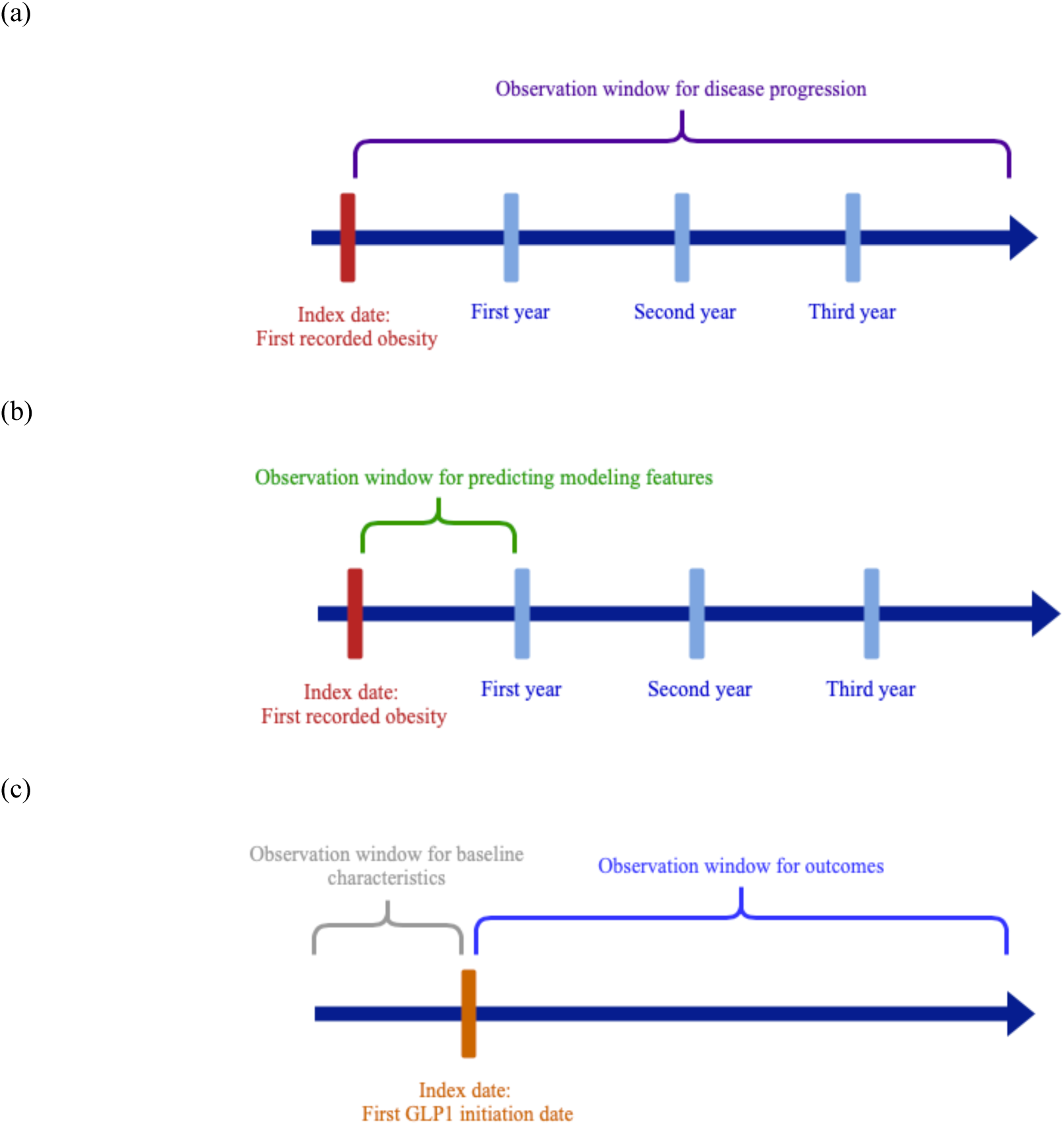
Study timeline for (a) obesity progression. (b) predicting modeling. (c) GLP-1RA heterogeneity treatment effect.

Then, we modeled the processed longitudinal EHRs as a disease progression graph (DPG). This graph structure effectively captures individual patient disease progression patterns and preserves interpatient progression correlations. Formally, we defined the DPG as a directed graph, 𝐷𝑃𝐺 = (𝒱, ℰ, 𝒜), where each node 𝑣 ∈ 𝒱 represents an enhanced encounter representation 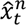. To construct the DPG, we began by identifying the top-k most similar neighbors for each node, utilizing a similarity function based on the Jaccard index.^21^ Subsequently, we inserted a directed edge 𝑒 ∈ ℰ between each pair of similar nodes, with the direction reflecting the chronological order of the encounters. In addition, we checked and inserted edges between nodes (i.e., enhanced encounter representation) from the same patient to ensure that the progression of the disease can be linked through a path. The adjacency matrix 𝒜 captures the weights of the edges (i.e., the elapsed time between two encounters). Figure 2b shows an example of the proposed DPG.

To generate outcome-oriented embeddings for each node, we developed a GNN-based encoder. This encoder utilizes the graph structures to propagate and aggregate node features in an iterative manner. The encoder contains two GNN layers and a fully connected layer to transform the learned embeddings to a specific label. The GNN layers^22^ can be described as:

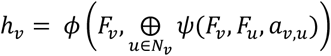

Where 𝐹 presents the features of a node and ℎ*_v_* refers to the learned embeddings for a node 𝑣, 𝜙 and 𝜓 are learnable functions, ⨁ is nonparametric operation (e.g., aggregation and concatenation), 𝑁_𝑣_ means the neighbors of 𝑣, and 𝑎*_v,u_* is the weight of edge between nodes 𝑣 and 𝑢. We utilized MagNet,^23^ a graph convolutional network (GCN) designed for directed graphs, as the core of the GNN layer. MagNet extends traditional GCNs by incorporating directional information via a complex Hermitian matrix. With its superior ability to encode structural information from directed graphs and outperforming traditional GCNs on various benchmarks, MagNet is an excellent candidate for modeling the intricate information of disease progression contained within DPGs. Additionally, we implemented three other GNN variants—GCN,^24^ graph attention network (GAT),^25^ and graph sample and aggregate network (GraphSAGE)^26^—to compare their performance in embedding learning. The embeddings for each node are generated by GNN layers, capturing the underlying disease progression information relevant to patient encounters. These outcome-oriented embeddings are then fed into the fully connected predictor, which forecasts the outcomes of the next encounter along the patient’s BMI change trajectory: decrease (≤−1), steady (−1 to 1), increase (1 to 5), or greatly increase (≥5). The fully connected layer can be formulated as:

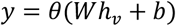

where 𝜃 is the activation function and 𝑊 and 𝑏 are the learnable parameters. During the training stage, we employed gradient descent to optimize the GNNs by minimizing the difference between the actual and predicted labels.

### Step 2: Identification of disease-progression subphenotypes via time-series clustering

After obtaining the learned embeddings of each node (i.e., enhanced encounter representation), we combined these embeddings into a sequence. Each patient had an embedding sequence arranged in chronological order based on the original longitudinal EHRs, described as 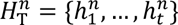, where 𝑛 and T are patient and time indexes, respectively. We then applied the time-series K-means clustering method with dynamic time warping (DTW)^27^ to identify similar latent characteristics among embedding sequences and determine progression subphenotypes. We chose time-series K-means due to its simplicity as an unsupervised algorithm, known for its rapid convergence, even on large datasets. Time-series K-means clusters similar time series data by grouping them based on their similarity. This iterative algorithm repeatedly reassigns time series to clusters and updates the centroids to minimize the within-cluster errors, as defined:

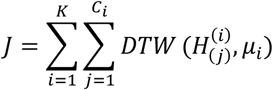

where 𝐾 is the number of clusters, 𝐶. refers to the number of patients in cluster 𝑖, 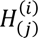 is the embedding sequence of the 𝑗-th patient in cluster 𝑖, and 𝜇. denotes the centroid of cluster 𝑖. The function DTW^27^ is a distance metric used to measure similarity between two embedding sequences of different lengths.

To determine the optimal 𝐾, we adopted a combination of quantitative and qualitative analyses. Specifically, we applied the time-series K-means algorithm with varying cluster numbers (K = 3 to 10) to generate clustering outcomes under different K settings. Then, we evaluated cluster quality using the Davies–Bouldin index (DBI)^28^ with DTW as the similarity measure; lower DBI indicates better clustering performance. After obtaining the candidate results (e.g., K from 3 to 4 in MagNet and GraphSAGE showing fair and acceptable quantitative performance), we examined the characteristics of each cluster under different settings and manually assessed cluster quality. This assessment considered factors including (1) the BMI Trajectory Group (BTG), a robust categorization designed to summarize longitudinal BMI change, and (2) selected obesity-related comorbidities reviewed by two reviewers with clinical backgrounds (J.G. and P.K) (**Table 1**). BTG comprises four categories: **substantial-increase**, **moderate**-**increase**, **steady**, and **decrease**. For each patient, annual BMI changes were first binned into four groups: ≤ −1, −1 to 1, 1 to 5, and ≥ 5 kg/m², encoded as 0, 1, 2, and 3, respectively. We then averaged these yearly codes over the observation window and assigned BTG as follows:

- **Substantial-Increase**: Average ≥ 1.5 with at least one “3”
- **Moderate-Increase**: Average ≥ 1.5 without any “3”
- **Steady**: Average 0.5 – <1.5
- **Decrease**: Average < 0.5

**Table 1.** Obesity related comorbidities.

| Category | Comorbidities |
| --- | --- |
| Cardiovascular Disorders (CVD)/MACE | Coronary Artery Disease, Heart Failure, Atrial Fibrillation |
| Metabolism & Nutrition (Endocrine) Disorder | Type 2 Diabetes CCW, Vitamin D deficiency, Gallbladder Disease, Acute/Chronic Pancreatitis |
| Gastrointestinal Disorders (GI) | GERD, Stroke/TIA, NAFLD/NASH |
| Mental Disorders | Depression |
| Renal & Urinary (kidney) Disorders | Chronic Kidney Disease (CKD) |
| Vascular Disorders | Pulmonary Embolism, Venous Thromboembolism |
| Others | Sleep apnea, Osteoarthritis |

GERD: Gastroesophageal reflux disease; TIA: Transient ischemic attack; NAFLD: Nonalcoholic fatty liver disease; NASH: Nonalcoholic steatohepatitis; MACE: Major adverse cardiovascular events.

This trajectory-based scheme integrates year-to-year variability, mitigates sensitivity to single measurements, and addresses the limitations of simple pre–post summaries, especially under unequal follow-up and irregular visit spacing. Two reviewers (YA.L. and J.G.) evaluated the cluster quality, and a consensus discussion, led by a third reviewer (Y.H.), was conducted to consolidate opinions and determine the optimal K.^29^ We then reran time series K-means with the optimal 𝐾 to classify the embedding sequences for each patient into distinct clusters, where each cluster describes a unique disease progression subphenotype.

### Step 3: Assessment of subphenotype interpretability by predictive modeling

To evaluate the clinical utility of the subphenotypes, we assessed their predictive validity using supervised models. Subphenotypes were first assigned to each patient by the proposed GNN framework. As illustrated in Figure 3b, we defined the index date as the patient’s first recorded obesity diagnosis and derived features from the subsequent 12 months (e.g., demographics, comorbidities, medications) to predict subphenotype membership. We implemented commonly used ML models for this purpose, including linear models (e.g., logistic regression^30^) and XGBoost.^31–36^ We also incorporated two imbalanced-data preprocessing methods: random oversampling and random undersampling. Finally, we utilized SHapley Additive exPlanations (SHAP),^37^ a widely used XAI technique, to identify important features contributing to the models’ ability to classify each patient’s subphenotype.

### Modeling procedures and benchmarks

We followed ML best practices: we stratified the data by patients and split it into training, validation, and testing sets according to a 70%:10%:20% ratio. We selected the Area under the Receiver Operating Characteristic Curve (AUROC) as our primary metric and included sensitivity, specificity, and precision as additional metrics for the GNNs and the subphenotype prediction model. Furthermore, we conducted a five-fold cross-validation Bayesian hyperparameter search on the training set to optimize the parameters of the subphenotype prediction models.

### Step 4: Heterogeneity treatment effect of GLP-1RAs across subphenotypes

Within each subphenotype, we emulated a target trial comparing initiation of GLP-1RA vs. non-users (Figure 3c). To mitigate confounding, we constructed the comparator group via time-conditional propensity score matching, pairing non-users to GLP-1RA initiators with index date aligned within **±**1 week of the initiator’s start date. We then fit Cox proportional hazards regression models to estimate hazard ratios (HRs) for each outcome, with 95% confidence intervals (CIs), comparing GLP-1RA initiators with matched non-initiators.

## Results

### Descriptive statistics of the study cohort

Our final analysis included 237,103 eligible patients with overweight or obesity. **Table 2** highlights the characteristics of the study cohort. The mean age of the cohort was 51.8 years (std = 17.3), and 58.1% were female. Patients were 48.4% non-Hispanic White (NHW), 27.9% non-Hispanic Black (NHB), and 13.0% Hispanic. The follow-up period ranged from 3.0 – 10.0 years, with an average of 4.9 years.

**Table 2.** Characteristics of the obesity progression study cohort.

|  | Study cohort (N = 237,103, patients diagnosed with obesity) |
| --- | --- |
| **Observation period, mean years (std)** | 4.91 (2.26) |
| **Age, mean (std)** | 51.78 (17.25) |
| **Female, N (%)** | 137,690 (58.07%) |
| **Race-ethnicity, N (%)** |  |
| Non-Hispanic White | 114,856 (48.44%) |
| Non-Hispanic Black | 66,242 (27.94%) |
| Hispanic | 30,778 (12.98%) |
| Others | 6,076 (2.56%) |
| Unknown | 19,151 (8.08%) |

### Identifying obesity progression subphenotypes

We identified three obesity progression subphenotypes using the proposed disease progression subphenotyping framework. Details of the framework optimization are reported in the supplement (**eResult 1**: model parameter tuning, and **eResult 2**: K-selection for the number of clusters), and the full experiments are shown in **e**Figures 2–7. Figure 4a shows the BTG distribution across subphenotypes. Patients in subphenotype 2 (S2) and S3 showed faster progression to obesity-related diseases, with a high percentage achieving substantial-increase BTG (43.6% and 33.4%, respectively). Conversely, individuals in S1 exhibited a more stable and slower progression to obesity-related diseases, with 64.4% classified as steady. Figure 4b visualizes the cumulative incidence of comorbidities and Kaplan-Meier curves, showing 10-year survival for each subphenotype after obesity diagnosis. There were notable differences (p < 0.001) among the various subphenotypes. S2 consistently demonstrated the highest cumulative incidence across most conditions, including coronary artery disease, HF, depression, T2D, and CKD, indicating the worst health trajectory. S1 generally showed the lowest cumulative incidence for most conditions, while S3 typically fell between the other two.

**Figure 4.**
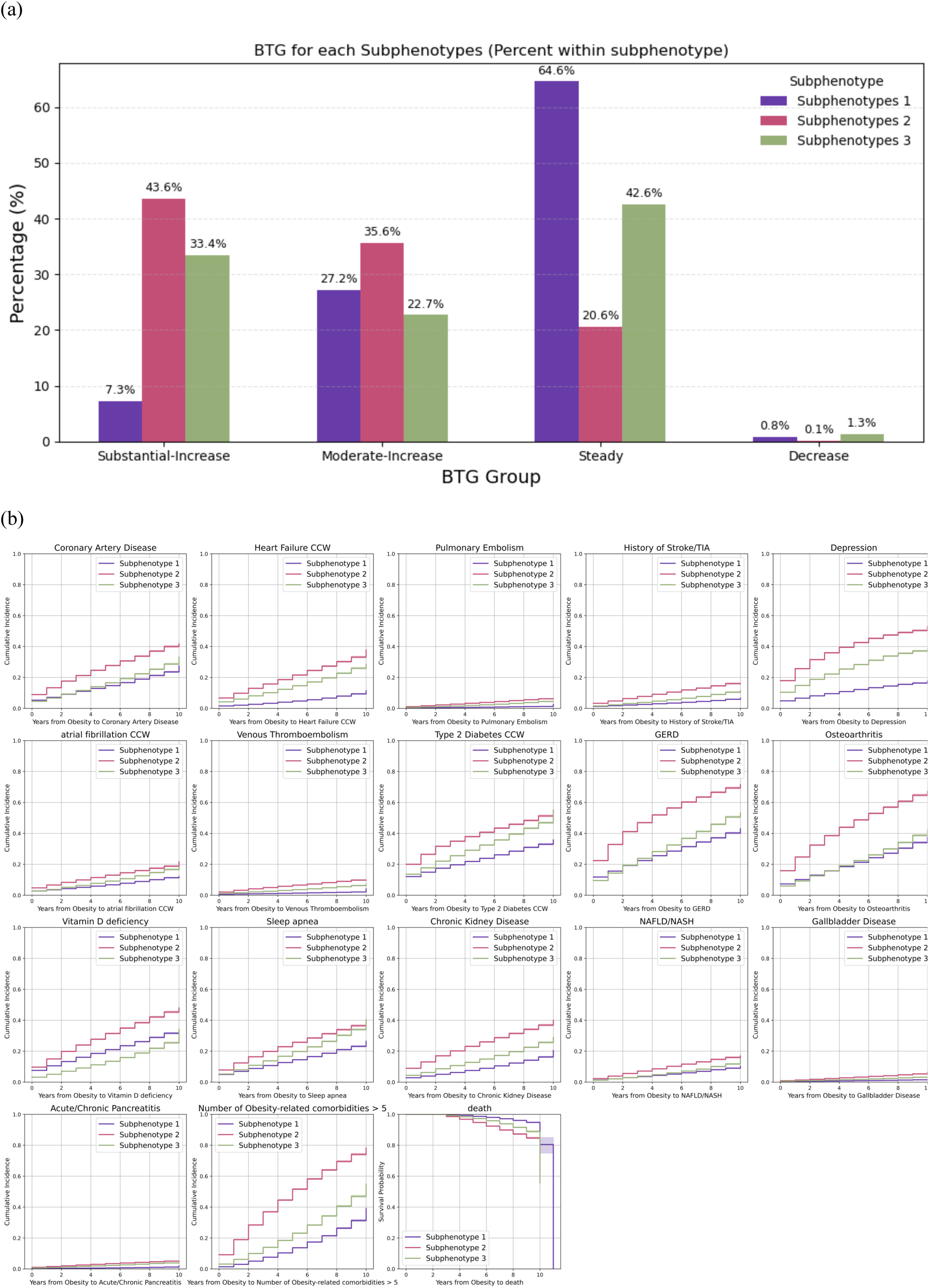
Characteristics of obesity progression subphenotypes identified by MagNet with time series K-means (K = 3). (a) BTG distribution in each subphenotype. (b) Cumulative incidence of obesity related comorbidities and Kaplan-Meier survival curves for each subphenotype.

**Figure 5.**
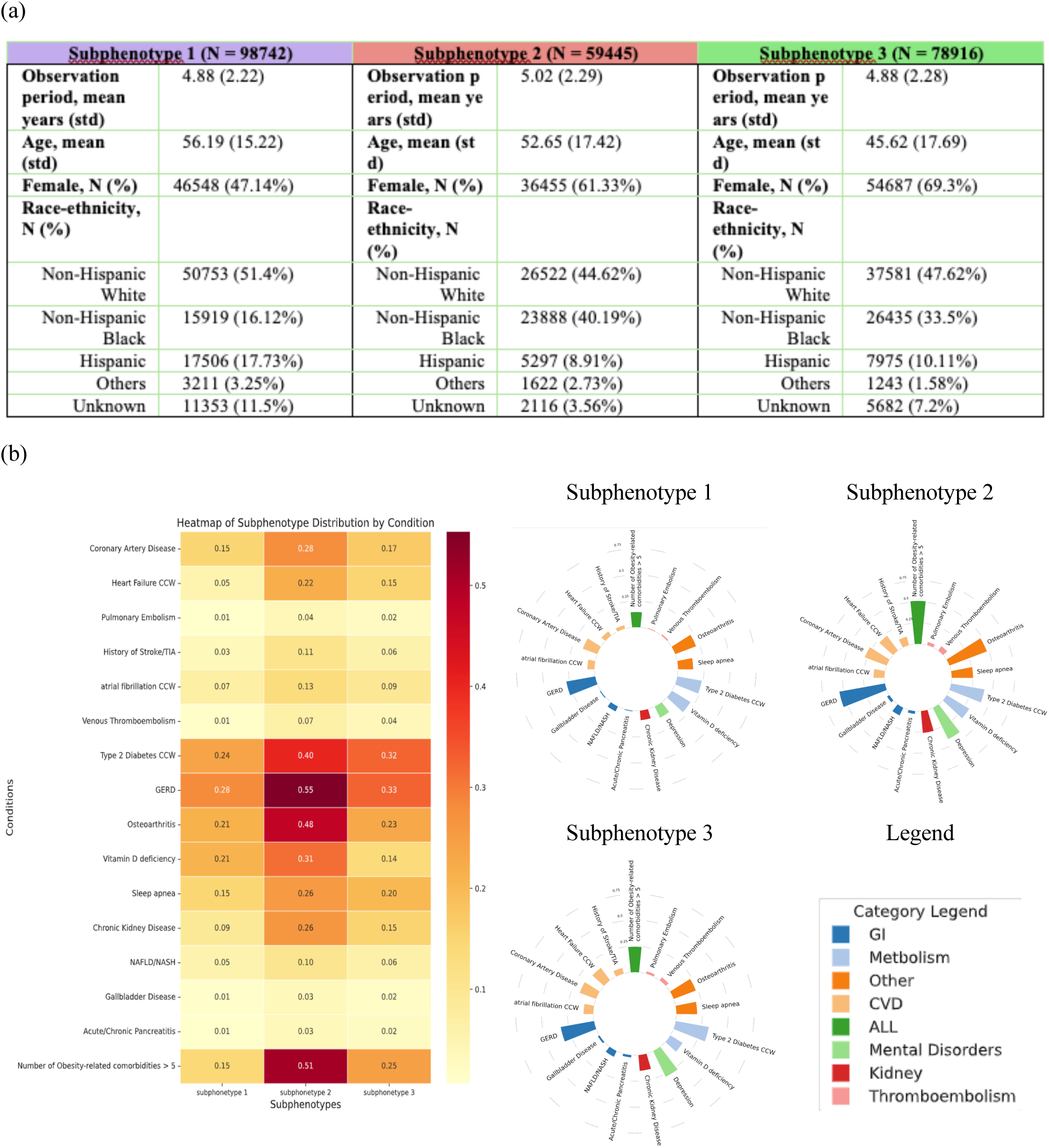
Characteristics of obesity progression subphenotypes identified by MagNet with time series K-means (K = 3). (a) Demographic characteristics of each subphenotype. (b) Heatmap and circle plot of the obesity related comorbidities prevalence across the subphenotypes.

**Figure 6.**
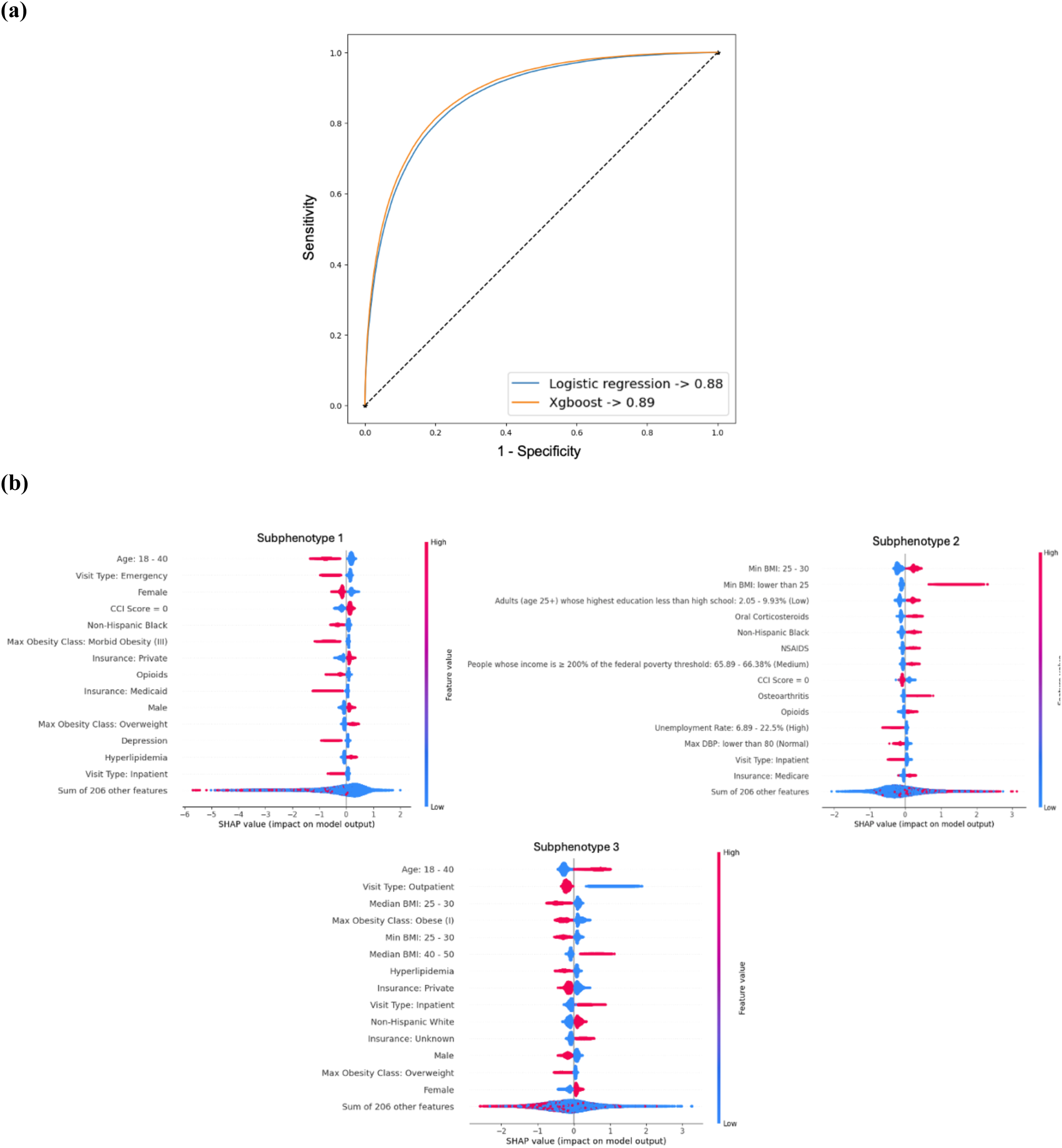
Comparative assessment of the predictability and interpretability of XGBoost and linear models in predicting obesity subphenotypes. (a) Model performance for predicting progression subphenotypes based on encounter information. (b) Analysis of feature importance in XGBoost using SHAP values.

**Figure 7.**
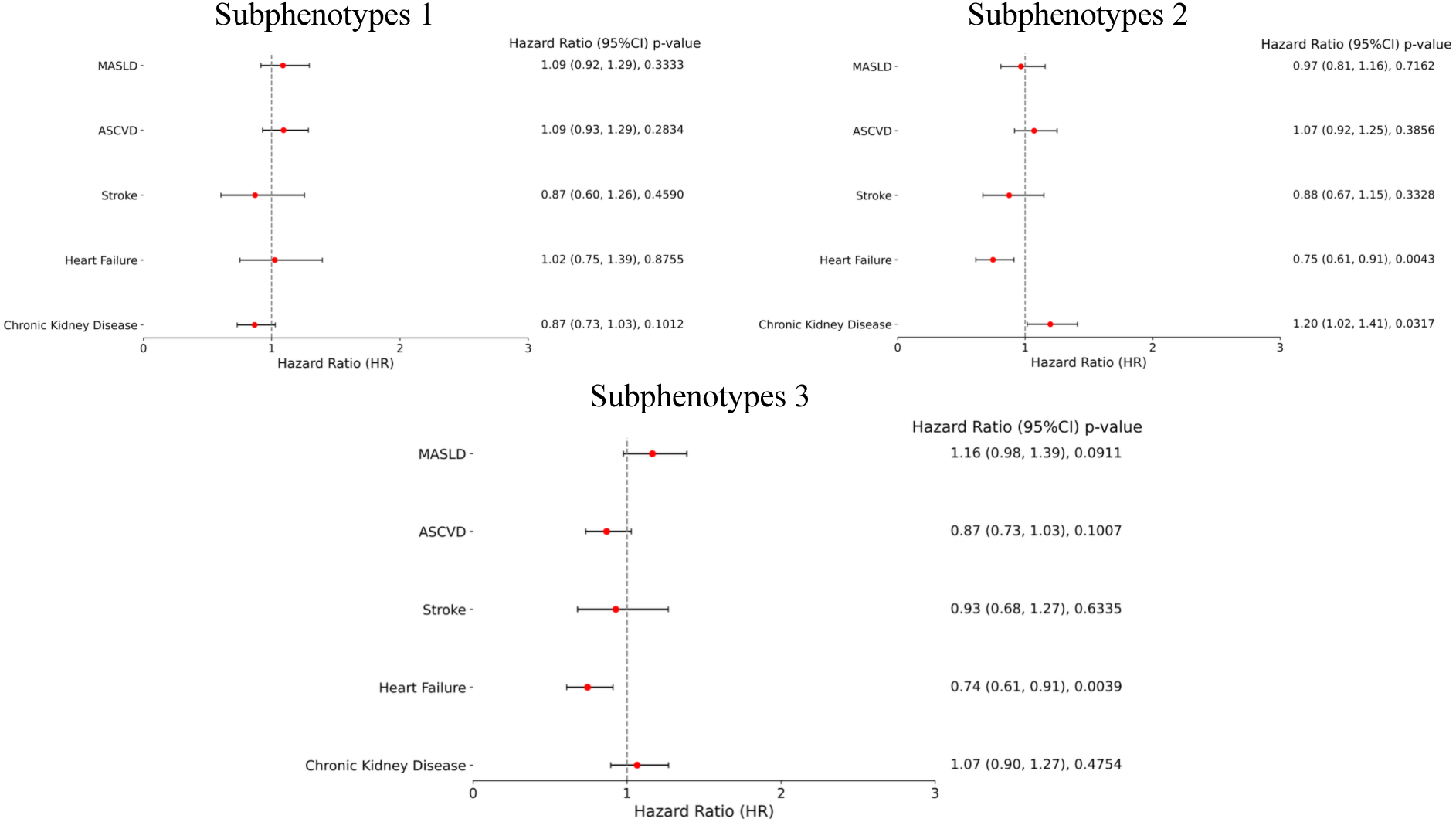
Hazard Ratios for GLP-1RA users vs non-users within each subphenotype identified by MagNet with time series K-means (K = 3). ASCVD: Metabolic dysfunction-associated steatotic liver disease; MASLD: Major atherosclerotic cardiovascular disease

Figure 5a presents demographic statistics for each subphenotype. Follow-up duration was comparable across subphenotypes (mean ≈4.9–5.0 years). S1 (N=98,742) had the highest mean age (56.2±15.2 years), a near-even sex distribution (47.1% female), and the highest proportion of NHW patients (51.4%). S2 (N=59,445) was moderately younger (52.6±17.4 years), majority female (61.3%), and had the largest share of NHB patients (40.2%). S3 (N=78,916) was the youngest (45.6±17.7 years) and predominantly female (69.3%), with substantial representation of NHB (33.5%) and NHW (47.6%) patients. Figure 5b summarizes the clinical profiles of the three subphenotypes. We focused on obesity-related comorbidities and quantified their prevalence within each group to identify distinguishing features. Gastroesophageal reflux disease (GERD) was the most prevalent condition across all subphenotypes. Pairwise comparisons using chi-square tests indicated significant differences in comorbidity prevalence between subphenotypes (all *p* < 0.001). Overall, S2 displayed the greatest multimorbidity burden, particularly for T2D, osteoarthritis, vitamin D deficiency, sleep apnea, and GERD, and included the largest share of patients with >5 obesity-related comorbidities. S3 showed intermediate prevalence, with relatively higher rates of GERD and T2D than S1, which generally exhibited the lowest prevalence across conditions. Taken together, these findings suggest a gradient of multimorbidity—S2 > S3 > S1—consistent with the observed BTG distribution.

### Predictability and interpretability of the identified subphenotypes

Figure 6a illustrates the performance of the prediction models, logistic regression and XGBoost, trained on one-year of post–index data without hyperparameter tuning or cross-validation and without any resampling. Performance was fair (AUROC 0.88–0.89), with XGBoost comparable to logistic regression. Full sensitivity analyses with tuning/cross-validation and oversampling/undersampling are reported in **e**Figure 8. Figure 6b presents SHAP values for the XGBoost model. Across subphenotypes, influential predictors included age, minimum BMI of the year (obesity class), encounter type, sex, race/ethnicity, use of gastrointestinal (GI)-risk analgesics (oral corticosteroids, nonsteroidal anti-inflammatory drugs (NSAIDs), opioids), and the Charlson Comorbidity Index. For S2 (progressive), higher predicted probability was associated with female sex, lower obesity class, and higher socioeconomic status (**e**Figure 9). However, socioeconomic status declined over follow-up in this group (**e**Figure 10), consistent with illness-related socioeconomic erosion^38^ as multimorbidity and treatment burden accumulate. We also observed higher exposure to corticosteroids, NSAIDs, and particularly opioids (**e**Figure 11), consistent with medication-related metabolic risk^39,40^ that can accelerate BMI increase and worsen comorbidity burden. Taken together, these findings suggest a trajectory in which initially better-resourced patients enter a progressive pathway that is subsequently reinforced by both socioeconomic deterioration and weight-promoting analgesic/steroid regimens.

### GLP-1RA treatment heterogeneity within each subphenotype

We then estimated subphenotype-specific hazard ratios (HRs) for GLP-1RA users vs non-users across five outcomes (Figure 7). In S1, associations were not statistically significant; HRs were near 1 for MASLD (HR 1.09; 95% CI, 0.92–1.29), ASCVD (1.09; 0.93–1.29), stroke (0.87; 0.60–1.26), HF (1.02; 0.75–1.39), and CKD (0.87; 0.73–1.03). In S2, estimates were null for MASLD (0.97; 0.81–1.16), ASCVD (1.07; 0.92–1.25), and stroke (0.88; 0.67–1.15); GLP-1RA use was associated with lower HF risk (0.75; 0.61–0.91; *p*=0.0043) but higher risk of CKD (1.20; 1.02–1.41; *p*=0.032). In S3, results were non-significant for MASLD (1.16; 0.98–1.39), ASCVD (0.87; 0.73–1.03), and stroke (0.93; 0.68–1.27); GLP-1RA use was associated with lower HF risk (0.74; 0.61–0.91; *p*=0.0039), with no difference for CKD (1.07; 0.90–1.27). Overall, GLP-1RA therapy demonstrated heterogeneous effects across subphenotypes, with a consistent reduction in HF risk in S2–S3, no clear benefit or harm for ASCVD or stroke, and a possible CKD risk increase confined to S2. Most other estimates clustered near the null. Cluster characteristics are reported in **eTable 6-8**.

## Discussion

In this study, we identified three obesity progression subphenotypes using large real-world EHR collections from the OneFlorida+ network, encompassing 237,103 patients with obesity and up to 10 years of observation. To achieve this, we developed a novel outcome-oriented GNN framework that models sequences of events in longitudinal EHRs as directed graphs and captures similar patient encounters through directed edges. This framework enables GNNs to generate representations for patients with similar characteristics while considering the changes in individual health conditions over time. Using time-series K-means clustering on the representations learned from GNNs, our approach can effectively model nuanced similarities in disease progression patterns. Our results indicate that this approach holds promise for detecting predictable obesity progression subphenotypes, providing valuable and explainable insights into the development of the disease.

Our identified subphenotypes with distinct obesity progression patterns suggest that obesity does not follow uniform transitions between disease states but rather exhibits heterogeneous progression pathways, aligning with the existing research.^41–44^ Several recent studies have used data-driven methods to characterize heterogeneity in obesity, but they differ in emphasis on progression versus clinical profiling. For example, Takeshita et al. identified seven subgroups of adults with obesity from population data using cluster analysis and showed prognostic differences across clusters (including a metabolically healthy group), but the work focused primarily on baseline complications and outcomes, not longitudinal BMI-change patterns.^41^ In contrast, multiple trajectory studies have modeled long-term BMI change, linking distinct BMI-trajectory groups to later cardiometabolic risk, but without detailed comorbidity-based subphenotyping of routine-care populations.^42,43^ Complementary EHR-based subphenotyping efforts have “deep-phenotyped” obesity cohorts and clustered patients using routinely collected data, highlighting multimorbidity structure, yet they stop short of defining progression subphenotypes anchored to BMI-change dynamics.^44^ Building on these strands, our framework derives outcome-oriented subphenotypes of obesity from routine-care records by (1) summarizing BMI-change trajectories, (2) pairing them with system-wide comorbidity profiles (e.g., cardiometabolic, gastrointestinal, and genitourinary), and (3) comparing rates of progression across subphenotypes. This approach bridges prior clustering that emphasized baseline subphenotypes or prognosis alone with trajectory analyses that emphasized BMI change without granular clinical context. In practice, such progression subphenotypes may help identify patients at higher risk of rapid worsening years before complications emerge—supporting earlier, targeted interventions and comorbidity management.

Beyond numerical differences, the three subphenotypes we identified outline actionable care pathways for obesity management. S2, marked by accelerated BMI gain and concentrated multimorbidity, likely reflects reinforcing loops: weight gain → pain/obstructive sleep apnea (OSA)/inactivity → further weight gain, amplified by social and medication factors. This pattern argues for early, intensive, multi-domain intervention: prompt anti-obesity pharmacotherapy when appropriate,^45^ systematic screening and treatment for OSA and T2D,^46,47^ pain-sparing strategies to reduce reliance on steroids/opioids/NSAIDs,^48^ and structured nutrition/physical activity programs that account for functional limitations.^49^ Health-system planning can prioritize this group for care escalation and closer follow-up, while reserving lower-touch maintenance approaches for the more stable subphenotype. S3 suggests a window for trajectory diversion; patients are not yet high burden but show signals (e.g., GERD, early metabolic disease) that respond to timely management. Embedding BTG into routine visits can trigger protocolized actions (e.g., HbA1c and OSA screening, medication review for weight-promoting drugs)^50^ before patients transition to the progressive group. S1 underscores that weight stability is clinically meaningful; preserving it, especially in older adults, may prevent downstream complications even without dramatic weight loss. Across all subphenotypes, socioeconomic status declined over follow-up; notably, S2 began with the most favorable SDOH profile yet exhibited the steepest erosion. This pattern suggests socioeconomic deterioration may both mediate and amplify rapid progression, warranting explicit modeling as a time-varying factor in future analyses and incorporation of routine SDOH monitoring and interventions in subphenotype-specific pathways. In practical terms, these subphenotypes can enable more targeted research and care. Trials can enroll the right patients for the right questions (e.g., test escalation vs. maintenance strategies within each subphenotype). Health systems can focus resources where they’re most needed (e.g., clinics or neighborhoods with many progressive patients) and address equity issues suggested by the demographic patterns. More broadly, pairing BMI trajectories with comorbidity profiles moves us beyond one-size-fits-all averages toward precision obesity care: matching treatment intensity, type, and follow-up frequency to a patient’s likely course.

In our analysis, the three subphenotypes derived by the algorithm exhibited distinct baseline risk profiles and divergent responses to the simulated GLP-1RA intervention, underscoring substantial biological heterogeneity. S2, with the highest incidence of cardiovascular, renal, and hepatic endpoints, showed a pronounced HF benefit from GLP-1RA (HR 0.75) but also a concerning signal of worsening CKD (HR 1.20). In contrast, S3 obtained clear benefit for HF with a neutral renal effect, while S1, characterized by lower baseline event rates, showed minimal measurable benefit. These patterns may be explained by the higher prevalence of moderate renal impairment (eGFR 30–60 mL observed in S2 (4.45%) compared with S1 (2.12%) and S3 (2.22%), suggesting that patients in S2 had reduced renal reserve at baseline, which may have made them more susceptible to hemodynamic changes associated with GLP-1RA therapy. Meanwhile, S3 may represent a “vascular-predominant” subphenotype with better preserved renal function, such that GLP-1RA’s metabolic and hemodynamic benefits translate to cardioprotection without renal penalty. These results align with meta-analytic evidence that GLP-1RAs confer cardioprotective effects broadly (e.g., reduced MACE) and modest kidney benefits, though effects on renal outcomes remain more inconsistent across baseline kidney function strata,^51^ and recent reviews emphasize that GLP-1RA benefits are often more robust for cardiovascular endpoints than for renal ones, especially in patients with diminished eGFR.^52^ Together, our findings argue for precision targeting of GLP-1RA interventions, where the same therapy may yield a net benefit in one subphenotype but require close renal monitoring or alternative strategies in another. Prospective validation is needed to confirm these differential responses and explore underlying mechanistic pathways.

Our study has several important clinical implications. First, the proposed framework refines obesity phenotyping by identifying distinct progression patterns that pair BMI trajectory patterns with comorbidity profiles, yielding a more precise view of the disease course than a single obesity label. Second, recognizing these progression patterns offers practical prognostic value for care planning; patients in the progressive group may warrant earlier escalation (e.g., GLP-1RA therapy when appropriate) and closer monitoring, whereas patients with stable trajectories may be suited to maintenance strategies. Finally, because subphenotypes may respond differently to treatment, testing subphenotype-specific responses supports more targeted and effective interventions and moves practice toward personalized obesity care.

Our study is subject to several limitations. First, the cohort was drawn largely from health systems in Florida, Georgia, and Alabama, which may limit generalizability despite substantial demographic diversity. Future work should validate the framework in other regions and networks. Second, while prediction models achieved fair discrimination, forecasting progression subphenotypes remains challenging when using routine EHR features alone; integrating knowledge-driven variables and richer longitudinal signals may improve accuracy. In addition, observational estimates (e.g., subphenotype-specific GLP-1RA effects) remain susceptible to residual confounding and incomplete capture of lifestyle and adherence; complementary sensitivity analyses and external prospective validation are needed to strengthen causal inference and assess transportability.

## Conclusion

Using a GNN framework that learns progression-aware representations coupled with time-series clustering, we identified three clinically distinct obesity progression subphenotypes from large-scale routine EHRs that differ in BMI trajectory, multimorbidity burden, and response to GLP-1RA initiation. Together, these findings provide interpretable insights into obesity progression that can inform the development of precision obesity care.

## Data Availability

The EHR data used in this study contain protected health information and cannot be shared publicly due to patient privacy and institutional policy. De-identified data may be made available to qualified researchers upon reasonable request and completion of a data use agreement and IRB approval from University of Florida. Requests should be directed through OneFlorida+ Clinical Research Network (email).

## Acknowledgements

This work was supported by the National Institutes of Health (NIH) (R01DK133465). The study was exempt approved by the University of Florida IRB (IRB202300903).

## Author Contributions

Conceptualization, J.G., J.B., and Y.H.; methodology, Y.H. and J.G.; formal analysis, YA.L; data curation, YA.L; resources, J.G.; writing – initial draft, YA.L., Y.H. and H.D.; critical review and editing, J.G., J.B., T.Y., and V.S.; supervision: J.G. and J.B.. All authors have read and agreed to the published version of the manuscript.

## Competing Interests

The authors declare no conflict of interest relevant to the study.

## Notes

### Competing Interest Statement

The authors have declared no competing interest.

### Author Declarations

The study was exempt approved by the University of Florida IRB (IRB202300903).

