## Supplemental Materials for "Characterize Disease Progression Subphenotypes in Real World Populations with Overweight and Obesity using a Graph-based Neural Network Framework"

This supplementary file contains the additional figures and tables of the experimental results for readers. The content is outlined as follows:


- **eResult 1.** GNN performance for learning disease progression representation
- **eResult 2.** DBI Screening for Optimal K
- **eTable 1.** Codes used for outcome definition and inclusion and exclusion criteria
- **eTable 2.** Variables used in study
- **eTable 3.** Exposures of interest
- **eTable 4.** Predictive performance of various GNNs (loss function = Cross-entropy) on the BMI change for patient follow-up encounters
- **eTable 5.** Predictive performance of various GNNs (loss function = Focal loss) on the BMI change for patient follow-up encounters
- **eTable 6.** GLP1 treatment effect baseline characteristics in subphenotype 1
- **eTable 7.** GLP1 treatment effect baseline characteristics in subphenotype 2
- **eTable 8.** GLP1 treatment effect baseline characteristics in subphenotype 3
- **eFigure 1.** Davies-Bouldin Index for time series k-means clustering (K = 2 to 10) utilizing embeddings from GraphSAGE and MagNet
- **eFigure 2.** Characteristics of obesity progression subphenotypes identified by GraphSAGE with time series K-means (K = 3). (a) BTG distribution in each subphenotype. (b) Cumulative incidence of obesity related comorbidities and Kaplan-Meier survival curves for each subphenotype
- **eFigure 3.** Characteristics of obesity progression subphenotypes identified by GraphSAGE with time series K-means (K = 3). (a) Demographic characteristics of each subphenotype. (b) Heatmap and circle plot of the obesity related comorbidities prevalence across the subphenotypes
- **eFigure 4.** Characteristics of obesity progression subphenotypes identified by GraphSAGE with time series K-means (K = 4). (a) BTG distribution in each subphenotype. (b) Cumulative incidence of obesity related comorbidities and Kaplan-Meier survival curves for each subphenotype
- **eFigure 5.** Characteristics of obesity progression subphenotypes identified by GraphSAGE with time series K-means (K = 4). (a) Demographic characteristics of each subphenotype. (b) Heatmap and circle plot of the obesity related comorbidities prevalence across the subphenotypes
- **eFigure 6.** Characteristics of obesity progression subphenotypes identified by MagNet with time series K-means (K = 4). (a) BTG distribution in each subphenotype. (b) Cumulative incidence of obesity related comorbidities and Kaplan-Meier survival curves for each subphenotype
- **eFigure 7.** Characteristics of obesity progression subphenotypes identified by MagNet with time series K-means (K = 4). (a) Demographic characteristics of each subphenotype. (b) Heatmap and circle plot of the obesity related comorbidities prevalence across the subphenotypes
- **eFigure 8.** Predicting obesity subphenotypes: XGBoost vs linear models
- **eFigure 9.** Socioeconomic status among subphenotypes in year 1

- **eFigure 10.** Socioeconomic status over follow-up period in (a) S1; (b) S2; (c) S3
- **eFigure 11.** Opioids usage among subphenotypes

**eResult 1. GNN performance for learning disease progression representation**

**eTables 4 to 5** present the performance analysis of four GNNs under different settings, including the choice of loss functions (e.g., cross-entropy or Focal loss[1]), intermediate node embedding size (e.g., 64 or 128) and the final embedding size (e.g., 64, 128 or 256) for learning outcome-oriented disease progression representations. Across all settings, performance was broadly comparable: all metrics varied by ≤0.02 across configurations, with no consistent front-runner. Increasing dimensionality yielded at most marginal gains, and focal loss provided minor, inconsistent differences relative to cross-entropy.

From **eTables 4 to 5**, we observed that MagNet and GraphSAGE achieved the strongest, with broadly comparable results. We therefore retained the best configuration for each:

- MagNet: # focal loss, intermediate embedding dimension = 128, output embedding dimension = 256
- GraphSAGE: # focal loss, intermediate embedding dimension = 128, output embedding dimension = 256

These models were then used to generate embeddings for the next phase of clustering experiments.

**eResult 2. DBI Screening for Optimal K**

**eFigure 1** presents the Davies-Bouldin Index (DBI) for time series k-means clustering (K = 2 to 10) using embeddings from GraphSAGE and MagNet. Quantitatively, only **GraphSAGE (K=2–3)** and **MagNet (K=2)** met our DBI criterion (<1). However, given the natural course of BMI change among patients with obesity, we required at least **three** progression types (increase, steady, decrease). Accordingly, we examined **K=3–4** solutions whose DBI values were close to 1 and profiled their characteristics.

**eTable 1. Codes used for outcome definition and inclusion and exclusion criteria**

| Outcome/Inclusion and exclusion criteria | ICD-9 | ICD-9-PCS | ICD-10 | ICD-10-PCS | CPT/HCPCS |
|---|---|---|---|---|---|
| Pregnancy | 630-679.x, V22-V24.x, V27, V30-V39.x, 765-766.x | 72, 73, 74, 69, 66.62 | O00-O9A.x, Z33, Z34, Z3A, Z37, Z38, P07 | 10xxxxx, 3E0xxx, 0UB5xxx,0UB6xxx, 0U7C7ZZ, 0W8NXZZ | 59000-59899, S0199, S2260-S2267 |
| Bariatric surgery | | 43.82, 43.89, 44.30, 44.31, 44.38, 44.39, 44.50, 44.68, 44.69, 44.95, 44.96, 44.97, 44.99, 45.51, 45.90 | | 0DB60Z3, 0DB63Z3, 0DB64Z3, 0DB67Z3, 0DB68Z3, 0D1647A, 0D164ZA, 0D168ZA | 43633, 43644, 43645, 43775, 43842, 43843, 43844, 43846, 43847, 43848 |
| ASCVD – Ischemic Heart disease | 410-414, V45.81, V45.82 | 00.66, 36.0-36.3, 36.9, 88.5, 37.8 | I20, I21, I22, I23, I24, I25 | | |
| ASCVD – Revascularization | | 00.66, 17.55, 36.01, 36.02, 36.05-36.07, 36.10-36.19 | | 0210xxx, 0211xxx, 0212xxx, 0270xxx | 92920, 92921, 92924, 92925, 92928, 92929, 92933, 92934, 92937, 92938, 92941, 92943, 92944, 92973, 92980-92984, 92995, 92996, 33510-33536, 33545, 33572 |
| | **ICD-9** | | **ICD-10** | | |
| ASCVD – Myocardial infarction | 410 | | I21 | | |
| ASCVD – Mortality | Directly identified through DEATH raw table | | | | |
| ASCVD – Unstable angina | 411.1 | | I20.0 | | |
| Active malignant neoplasm | 140 – 208.xx | | C00–C96.x, C76–C80.x | | |
| MASLD – Steatotic | 571.8, 571.9 | | K76.0, K75.81 | | |
| MASLD - Hepatocellular carcinoma | 155.0 | | C22.0, C22.8 | | |
| MASLD – Liver cirrhosis | 571.2, 571.5, 572.3, 456.1, 456.21 | | K70.30, K74.60, K74.69, K76.6, I85.00, I85.10 | | |
| MASLD – Hepatic decompensation | 789.5, 789.59, 567.23, 456.0, 456.2, 456.8, 572.2, 572.4 | | R18.8, K70.31, K65.2, I85.01, I85.11, I86.4, K76.82, K72.91, K72.01, K72.11, K76.745 | | |
| Chronic Kidney Disease | 585 | | N18 | | |
| Stroke/TIA | 430, 431, 433.x1, 434, 435, 436, 997.02 | | I60, I61, I63, I66, G46, I97.81/I97.82, G97.3 | | |
| Heart Failure | 398.91, 402.x1, 404.x1, 404.x3, 428 | | I09.81, I11.0, I13.0, I13.2, I50 | | |

ASCVD: Metabolic dysfunction-associated steatotic liver disease; MASLD: Major atherosclerotic cardiovascular disease; TIA: Transient ischemic attack.

**eTable 2. Variables used in study**

| Variables Type | Variables name |
|---|---|
| Social determinants of health (SDOH) | AHRF_UNEMPLOYED_RATE (unemployment rate): Unemployment rate per 100 population (ages 16 and over) |
| | ACS_PCT_PERSON_INC_ABOVE200 (income level): Percentage of population with an income to poverty ratio of 2.00 or higher |
| | ACS_MEDIAN_HH_INC (median house income): Median household income (dollars, inflation-adjusted to data file year) |
| | ACS_PCT_LT_HS (education level): Percentage of population with less than high school education (ages 25 and over) (lower the value, higher the education level) |
| | ACS_PCT_HH_FOOD_STMP(community food access level): Percentage of households that received food stamps/SNAP, past 12 months |
| Demographics | Age, Sex, Race_ethnicity (NHW, NHB, Hispanic, Others, Unknown), Inpatient/outpatient visit, Insurance coverage (Medicare, Medicaid, No payment, Other Othergov, Private, Unknown) |
| Vitals | BMI, Systolic blood pressure, Diastolic blood pressure |
| Labs | Hemoglobin A1c, Total cholesterol, High-density lipoprotein (HDL), Low-density lipoprotein (LDL), estimated glomerular filtration rate (eGFR) |
| Comorbidities | Pulmonary embolism, Atrial fibrillation, Venous thromboembolism, Hypertension, Hyperlipidemia, Type 2 diabetes, GERD, Depression, Osteoarthritis, Vitamin D deficiency, Sleep apnea, prediabetes, Gallbladder disease, Metabolic syndrome, dyspepsia, Inflammatory bowel disease, Acute/Chronic pancreatitis, Anorexia, Cushing syndrome, Feeding difficulties, Congestive heart failure, Peripheral vascular disease, Cerebrovascular disease, Dementia, Chronic pulmonary disease, Rheumatic disease, Peptic ulcer disease, Mild liver disease, Diabetes without chronic complication, Diabetes with chronic complication, Hemiplegia or paraplegia, Renal disease, AIDS/HIV, Deep vein thrombosis (DVT), Anxiety, Sleep disorder, Apathy, Hypothyroidism, Hyperthyroidism, Seizures, PTSD, Bipolar disorder, Schizophrenia, OCD, Arthritis, Asthma, Hearing impairment, Vision impairment, Significant alcohol use, CCI score |
| Medications | Phentermine/Topiramate, Bupropion/Naltrexone, Lorcaserin, Anorectic drugs, SGLT2, Sulfonylurea, CCB, Beta blockers, Statin, Insulin, ACEIs, ARBs, Antidepressants, diuretic, Metformin, Thiazolidinedione, DPP4i, NSAIDS, Antipsychotics, Alpha glucosidase inhibitor, Meglitinide, Anti-dementia, Anticoagulant, Antiplatelets, Aldosterone receptor antagonists, Opioids, Oral corticosteroids, Orlistat |

DPP4i, dipeptidyl peptidase 4 inhibitors; SGLT2 Inhibitors, sodium-glucose cotransporter 2 inhibitors; ACEI, angiotensin-converting-enzyme inhibitors; ARB, angiotensin II receptor blockers; NSAIDS, non-steroidal anti-inflammatory drugs; CCB, Calcium channel blockers; GERD, gastroesophageal reflux disease; NAFLD, nonalcoholic fatty liver disease; NASH, nonalcoholic steatohepatitis; OCD, Obsessive-compulsive disorder; PTSD, Post-traumatic stress disorder; CCI, Carlson comorbidity index; Others: include American Indian or Alaska Native, Asian, Native Hawaiian or Other Pacific Islander, and individuals identifying as multiracial.

**eTable 3**. **Exposures of interest**

| | | |
|---|---|---|
| **GLP1RAs (GLP1)** | Liraglutide (LIRA): | **NDC**: 00169280015, 00169280090, 00169280097, 50090425700, 00169406012, 00169406013, 00169406090, 00169406099, 50090285300, 50090450300, 00169406097, 00169406098, 00169291115<br>**RxNorm**: 1598264,1598268,1598267,1598265, 897120, 1163230, 475968, 897124, 897126, 1186578, 897123, 897120, 897122, 1163230, 1727493, 897122 |
| | Semaglutide (SEMA): | **NDC**: 00169450101, 00169450114, 00169450501, 00169450514, 00169451701, 00169451714, 00169452401, 00169452414, 00169452501, 00169452514, 00169452590, 00169452594, 00169413602, 00169477212, 00169477297, 50090605100, 50090594900, 00169413212, 00169413297, 00169418113, 00169418197, 00169413013, 00169413001, 00169413211, 00169413290, 00169413611, 00169418190, 00169477211, 00169477290, 00169418103, 00169430301, 00169430313, 00169430390, 00169430393, 00169430330, 00169430399, 00169431401, 00169431413, 00169431430, 00169430701, 00169430713, 00169430730<br>**RxNorm**: 2553503, 2553602, 2553608, 2553902, 2554103, 2553506, 2553603, 2553803, 2553903, 2554104, 2554101, 2553501, 2553601, 2553802, 2553901, 2554102, 2553505, 2553502, 1991302, 1991304, 2619153, 1991308, 2599364, 1991311, 1991317, 2619154, 2398842, 2599365, 1991306, 2619152, 1991316, 2398841, 2599362, 1991310, 1991307, 1991302, 1991304, 2200653, 2200657, 2200646',2200654, 2200658, 2200650, 2200652, 2200655, 2200640, 2200644, 2200656, 2200645, 2200641, 2200644 |
| | Tirzepatide (TIRZ): | **NDC**: 00002150661, 00002145780, 00002146080, 00002147180, 00002148480, 00002149580, 00002150680, 00002115201, 00002124301, 00002145701, 00002146001, 00002147101, 00002148401, 00002149501, 00002150601, 00002221401, 00002234001, 00002242301, 00002300201, 00002015201, 00002024301, 00002121401, 00002134001, 00002142301, 00002200201, 00002245701, 00002245780, 00002246001, 00002246080, 00002247101, 00002247180, 00002248401, 00002248480, 00002249501, 00002249580, 00002250601, 00002250661, 00002250680<br>**RxNorm**: 2644413, 2644409, 2644415, 2644417, 2601736, 2601745, 2601763, 2601781, 2601769, 2601775, 2601757, 2601764, 2644407, 2601746, 2644403, 2601785, 2644399, 2601770, 2644419, 2601776, 2601758, 2644411, 2601754, 2601772, 2601766, 2601778, 2601742, 2601760, 2601761, 2644405, 2601743, 2644401, 2601784, 2601767, 2601773, 2601755, 2601730, 2601723, 2669704, 2669721, 2669718, 2669715, 2669712, 2669709, 2669706, 2669720, 2669717, 2669714, 2669711, 2669708, 2669703, 2669702, 266970, 2669709, 2601754, 2601772, 2601766, 2601778, 2601742, 2601760, 2601761, 2644405, 2601743, 2644401, 2601784, 2601767, 2601730, 2601723 |
| | Lixisenatide (LIXI): | **NDC**: 00024576101, 00024576105, 00024574502, 00024574702, 00024574502, 00024574101, 00024574000, 00024574702, 00024576101, 00024576102, 00024576105, 00024576302<br>**RxNorm**: 1440051, 1858994, 1440051, 1858994, 1803887, 1858996, 1803903, 1803902, 1440051, 1858994, 1803893, 1803896, 1859000, 1803888, 1803895, 1858997, 18038 |

| | | |
|---|---|---|
| | | 89, 1858998, 1803892, 1803894, 1858995, 1440052, 1440056, 1858991, 1803885, 1858993, 1440053, 1858992, 1803890, 1858999 |
| | Albiglutide (ALBI): | **NDC**: 00173086601, 00173086602, 00173086635, 00173086661, 00173086701, 00173086702, 00173086735, 00173086761<br>**RxNorm**: 1534801, 1534763, 1534805, 1534822, 1534802, 1534821, 1659117, 1534800, 1534820, 1534797, 1534819, 1659115, 1534798, 1534804 |
| | Exenatide (EXEN): | **NDC**: 00310653004, 00310654004, 00310654085, 00310651201, 00310652401, 00310651201, 00310652401, 00002021008, 00310652401, 54868538401, 66029021008, 66780021008, 66780021201, 66914103505, 68258894802, 00002021007, 00002021009, 00310651201, 00310651285, 54868538400, 54868538402, 66029021007, 66780021007, 66780021009, 66914103504, 68258894701, 00310654001, 00310654004, 00310654085, 00310653001, 00310653004, 00310653085, 00310652004, 66780021902, 66780021904, 66780022601<br>**RxNorm**: 60548, 60548, 604751, 1242964, 60548, 847913, 847917, 1990869, 1544918, 1242968, 847911, 847916, 1990867, 1653613, 1242965, 1990868, 1653625, 1653614, 1653619, 1990866, 847910, 847915, 1242963, 1544916, 1990864, 847908, 847914, 1653610, 1242961, 1990865, 1653611, 1653616, 1163790, 1169415, 1242967 |
| | Dulaglutide (DULA): | **NDC**: 00002143361, 00002143380, 00002143461, 00002143480, 00002223662, 00002318262, 50090546701, 50090348401, 50090348301, 00002223681, 00002318281, 00002143301, 00002143361, 00002143380, 50090348400, 54568043363, 54568043371, 00002143401, 00002143461, 00002143480, 50090348300, 54568043463, 54568043471, 00002223601, 00002223661, 00002223680, 50090546700, 00002318201, 00002318261, 00002318280<br>**RxNorm**: 1551291, 1551296, 1551291, 1551300, 1551306, 2395779, 2395785, 1551297, 1551305, 2395778, 2395784, 1649586, 1551295, 1551304, 2395777, 2395783, 1551292, 1551303, 2395776, 2395782, 1649584, 1551293, 1551299 |

**eTable 4. Predictive performance of various GNNs (loss function = Cross-entropy) on the BMI change for patient follow-up encounters**

| | Model | Accuracy | F1 | Precision | Recall | AUROC | Specificity |
|---|---|---|---|---|---|---|---|
| **Node dim = 64**<br>**Output dim = 64** | MagNet | 0.5145 | 0.3223 | 0.5399 | 0.3317 | 0.5613 | 0.7909 |
| | GCN | 0.5124 | 0.3259 | 0.5164 | 0.3321 | 0.5606 | 0.7890 |
| | GAT | 0.5099 | 0.3449 | 0.4853 | 0.3455 | 0.5693 | 0.7872 |
| | GraphSAGE | 0.5194 | 0.3450 | 0.5439 | 0.3456 | 0.5701 | 0.7945 |
| **Node dim = 64**<br>**Output dim = 128** | MagNet | 0.5148 | 0.3326 | 0.5307 | 0.3377 | 0.5643 | 0.7909 |
| | GCN | 0.5122 | 0.3258 | 0.5141 | 0.3316 | 0.5602 | 0.7888 |
| | GAT | 0.5115 | 0.3440 | 0.4840 | 0.3430 | 0.5679 | 0.7928 |
| | GraphSAGE | 0.5190 | 0.3430 | 0.5426 | 0.3439 | 0.5689 | 0.7939 |
| **Node dim = 64**<br>**Output dim = 256** | MagNet | 0.5147 | 0.3438 | 0.5101 | 0.3455 | 0.5683 | 0.7912 |
| | GCN | 0.5121 | 0.3241 | 0.5202 | 0.3313 | 0.5599 | 0.7886 |
| | GAT | 0.5111 | 0.3441 | 0.4774 | 0.3422 | 0.5671 | 0.7921 |
| | GraphSAGE | 0.5180 | 0.3395 | 0.5433 | 0.3415 | 0.5670 | 0.7925 |
| **Node dim = 128**<br>**Output dim = 64** | MagNet | 0.5166 | 0.3586 | 0.5018 | 0.3550 | 0.5740 | 0.7930 |
| | GCN | 0.5162 | 0.3407 | 0.5170 | 0.3419 | 0.5670 | 0.7920 |
| | GAT | 0.5110 | 0.3458 | 0.4765 | 0.3435 | 0.5677 | 0.7920 |
| | GraphSAGE | 0.5205 | 0.3496 | 0.5359 | 0.3481 | 0.5712 | 0.7944 |
| **Node dim = 128**<br>**Output dim = 128** | MagNet | 0.5162 | 0.3526 | 0.5055 | 0.3507 | 0.5716 | 0.7925 |
| | GCN | 0.5109 | 0.3290 | 0.5149 | 0.3333 | 0.5603 | 0.7874 |
| | GAT | 0.5073 | 0.3487 | 0.4772 | 0.3473 | 0.5707 | 0.7940 |
| | GraphSAGE | 0.5208 | 0.3537 | 0.5291 | 0.3510 | 0.5731 | 0.7951 |
| **Node dim = 128**<br>**Output dim = 256** | MagNet | 0.5188 | 0.3509 | 0.5204 | 0.3492 | 0.5715 | 0.7938 |
| | GCN | 0.5139 | 0.3356 | 0.5180 | 0.3380 | 0.5641 | 0.7902 |
| | GAT | 0.5084 | 0.3329 | 0.4894 | 0.3372 | 0.5651 | 0.7931 |
| | GraphSAGE | 0.5205 | 0.3556 | 0.5241 | 0.3521 | 0.5734 | 0.7947 |

**eTable 5. Predictive performance of various GNNs (loss function = Focal loss) on the BMI change for patient follow-up encounters**

| | Model | Accuracy | F1 | Precision | Recall | AUROC | Specificity |
|---|---|---|---|---|---|---|---|
| **Node dim = 64**<br>**Output dim = 64** | MagNet | 0.5124 | 0.3440 | 0.4956 | 0.3455 | 0.5682 | 0.7910 |
| | GCN | 0.5127 | 0.3334 | 0.5094 | 0.3377 | 0.5636 | 0.7895 |
| | GAT | 0.5087 | 0.3395 | 0.4690 | 0.3481 | 0.5627 | 0.7952 |
| | GraphSAGE | 0.5171 | 0.3414 | 0.5415 | 0.3428 | 0.5683 | 0.7937 |
| **Node dim = 64**<br>**Output dim = 128** | MagNet | 0.5134 | 0.3384 | 0.5117 | 0.3403 | 0.5657 | 0.7910 |
| | GCN | 0.5135 | 0.3351 | 0.5140 | 0.3385 | 0.5648 | 0.7911 |
| | GAT | 0.5091 | 0.3472 | 0.4721 | 0.3453 | 0.5689 | 0.7925 |
| | GraphSAGE | 0.5163 | 0.3388 | 0.5399 | 0.3415 | 0.5674 | 0.7933 |
| **Node dim = 64**<br>**Output dim = 256** | MagNet | 0.5129 | 0.3399 | 0.4983 | 0.3418 | 0.5664 | 0.7910 |
| | GCN | 0.5111 | 0.3252 | 0.5098 | 0.3310 | 0.5599 | 0.7889 |
| | GAT | 0.5101 | 0.3528 | 0.4719 | 0.3488 | 0.5712 | 0.7937 |
| | GraphSAGE | 0.5168 | 0.3405 | 0.5363 | 0.3423 | 0.5680 | 0.7938 |
| **Node dim = 128**<br>**Output dim = 64** | MagNet | 0.5157 | 0.3468 | 0.5092 | 0.3471 | 0.5701 | 0.7931 |
| | GCN | 0.5070 | 0.3418 | 0.5050 | 0.3497 | 0.5621 | 0.7946 |
| | GAT | 0.5073 | 0.3485 | 0.4734 | 0.3453 | 0.5695 | 0.7937 |
| | GraphSAGE | 0.5190 | 0.3501 | 0.5315 | 0.3484 | 0.5718 | 0.7952 |
| **Node dim = 128**<br>**Output dim = 128** | MagNet | 0.5157 | 0.3520 | 0.5003 | 0.3492 | 0.5715 | 0.7938 |
| | GCN | 0.5062 | 0.3474 | 0.5076 | 0.3459 | 0.5694 | 0.7928 |
| | GAT | 0.5094 | 0.3518 | 0.4858 | 0.3451 | 0.5749 | 0.7946 |
| | GraphSAGE | 0.5193 | 0.3512 | 0.5291 | 0.3491 | 0.5718 | 0.7945 |
| **Node dim = 128**<br>**Output dim = 256** | MagNet | 0.5159 | 0.3538 | 0.5019 | 0.3518 | 0.5731 | 0.7944 |
| | GCN | 0.5069 | 0.3528 | 0.4976 | 0.3563 | 0.5658 | 0.7952 |
| | GAT | 0.5051 | 0.3373 | 0.4822 | 0.3396 | 0.5663 | 0.7930 |
| | GraphSAGE | 0.5197 | 0.3553 | 0.5229 | 0.3518 | 0.5733 | 0.7949 |

**eTable 6. GLP1-RAs treatment effect baseline characteristics in subphenotype 1**

| Variable | Users, N (%) | Non-Users, N (%) | SEMA_user, N (%) | LIRA_user, N (%) | TIRZ_user, N (%) |
|---|---|---|---|---|---|
| Total | 3537 (50.0)% | 3537 (50.0)% | 2456 (69.44)% | 1118 (31.61)% | 522 (14.76)% |
| AGE (mean) | 54.66 | 55.89 | 54.73 | 54.18 | 53.75 |
| **Vital** | | | | | |
| BMI (mean) | 33.94 | 34.07 | 34.03 | 33.97 | 34.44 |
| Systolic blood pressure (mean) | 132.29 | 132.78 | 132.47 | 131.97 | 132.31 |
| Diastolic blood pressure (mean) | 80.21 | 80.43 | 80.32 | 79.7 | 80.67 |
| **Race Ethnicity** | | | | | |
| Hispanic | 832 (23.52)% | 662 (18.72)% | 639 (26.02)% | 226 (20.21)% | 137 (26.25)% |
| NHB | 576 (16.28)% | 658 (18.6)% | 414 (16.86)% | 185 (16.55)% | 62 (11.88)% |
| NHW | 1578 (44.61)% | 1648 (46.59)% | 996 (40.55)% | 565 (50.54)% | 228 (43.68)% |
| Others | 156 (4.41)% | 147 (4.16)% | 114 (4.64)% | 46 (4.11)% | 28 (5.36)% |
| Unknown | 395 (11.17)% | 422 (11.93)% | 293 (11.93)% | 96 (8.59)% | 67 (12.84)% |
| **Sex** | | | | | |
| Female | 2094 (59.2)% | 2122 (59.99)% | 1440 (58.63)% | 638 (57.07)% | 337 (64.56)% |
| Male | 1443 (40.8)% | 1415 (40.01)% | 1016 (41.37)% | 480 (42.93)% | 185 (35.44)% |
| **Comorbidities** | | | | | |
| Pulmonary Embolism | 8 (0.23)% | 8 (0.23)% | 5 (0.2)% | 2 (0.18)% | 1 (0.19)% |
| atrial fibrillation CCW | 48 (1.36)% | 77 (2.18)% | 38 (1.55)% | 6 (0.54)% | 8 (1.53)% |
| Venous Thromboembolism | 21 (0.59)% | 19 (0.54)% | 12 (0.49)% | 7 (0.63)% | 1 (0.19)% |
| Hypertension | 2158 (61.01)% | 2333 (65.96)% | 1568 (63.84)% | 602 (53.85)% | 320 (61.3)% |
| Hyperlipidemia CCW | 2283 (64.55)% | 2422 (68.48)% | 1692 (68.89)% | 583 (52.15)% | 360 (68.97)% |
| Type 2 Diabetes CCW | 2148 (60.73)% | 2135 (60.36)% | 1555 (63.31)% | 700 (62.61)% | 307 (58.81)% |
| GERD | 777 (21.97)% | 912 (25.78)% | 567 (23.09)% | 174 (15.56)% | 131 (25.1)% |
| Depression | 430 (12.16)% | 532 (15.04)% | 318 (12.95)% | 96 (8.59)% | 72 (13.79)% |
| Osteoarthritis | 516 (14.59)% | 665 (18.8)% | 391 (15.92)% | 110 (9.84)% | 81 (15.52)% |
| Vitamin D deficiency | 1035 (29.26)% | 1106 (31.27)% | 777 (31.64)% | 226 (20.21)% | 183 (35.06)% |
| Sleep apnea | 501 (14.16)% | 568 (16.06)% | 379 (15.43)% | 105 (9.39)% | 91 (17.43)% |
| prediabetes | 1042 (29.46)% | 1289 (36.44)% | 791 (32.21)% | 178 (15.92)% | 203 (38.89)% |
| Gallbladder Disease | 14 (0.4)% | 21 (0.59)% | 10 (0.41)% | 4 (0.36)% | 2 (0.38)% |
| Metabolic Syndrome | 95 (2.69)% | 94 (2.66)% | 77 (3.14)% | 16 (1.43)% | 19 (3.64)% |
| Dyspepsia | 18 (0.51)% | 23 (0.65)% | 13 (0.53)% | 7 (0.63)% | 1 (0.19)% |
| Inflammatory Bowel Disease | 25 (0.71)% | 32 (0.9)% | 14 (0.57)% | 7 (0.63)% | 6 (1.15)% |
| Acute/Chronic Pancreatitis | 13 (0.37)% | 35 (0.99)% | 9 (0.37)% | 5 (0.45)% | 0 (0.0)% |
| Anorexia | 6 (0.17)% | 4 (0.11)% | 4 (0.16)% | 1 (0.09)% | 2 (0.38)% |
| Cushing Syndrome | 4 (0.11)% | 5 (0.14)% | 4 (0.16)% | 0 (0.0)% | 0 (0.0)% |
| Feeding Difficulties | 390 (11.03)% | 429 (12.13)% | 310 (12.62)% | 50 (4.47)% | 83 (15.9)% |
| Congestive heart failure | 8 (0.23)% | 9 (0.25)% | 4 (0.16)% | 4 (0.36)% | 2 (0.38)% |
| Peripheral vascular disease | 86 (2.43)% | 111 (3.14)% | 70 (2.85)% | 19 (1.7)% | 10 (1.92)% |
| Cerebrovascular disease | 35 (0.99)% | 58 (1.64)% | 29 (1.18)% | 2 (0.18)% | 5 (0.96)% |
| Dementia | 3 (0.08)% | 6 (0.17)% | 1 (0.04)% | 1 (0.09)% | 0 (0.0)% |
| Chronic pulmonary disease | 348 (9.84)% | 411 (11.62)% | 266 (10.83)% | 65 (5.81)% | 62 (11.88)% |
| Rheumatic disease | 68 (1.92)% | 79 (2.23)% | 47 (1.91)% | 14 (1.25)% | 10 (1.92)% |
| Peptic ulcer disease | 19 (0.54)% | 21 (0.59)% | 11 (0.45)% | 3 (0.27)% | 5 (0.96)% |
| Mild liver disease | 47 (1.33)% | 63 (1.78)% | 32 (1.3)% | 14 (1.25)% | 9 (1.72)% |
| Diabetes without chronic complication | 2161 (61.1)% | 2146 (60.67)% | 1564 (63.68)% | 701 (62.7)% | 310 (59.39)% |
| Diabetes with chronic complication | 492 (13.91)% | 496 (14.02)% | 354 (14.41)% | 167 (14.94)% | 62 (11.88)% |
| Hemiplegia or paraplegia | 7 (0.2)% | 6 (0.17)% | 4 (0.16)% | 3 (0.27)% | 0 (0.0)% |
| Renal disease | 7 (0.2)% | 4 (0.11)% | 4 (0.16)% | 2 (0.18)% | 0 (0.0)% |
| AIDS/HIV | 13 (0.37)% | 17 (0.48)% | 11 (0.45)% | 3 (0.27)% | 3 (0.57)% |
| Deep Vein Thrombosis (DVT) | 11 (0.31)% | 11 (0.31)% | 6 (0.24)% | 4 (0.36)% | 0 (0.0)% |
| Anxiety | 618 (17.47)% | 713 (20.16)% | 446 (18.16)% | 115 (10.29)% | 122 (23.37)% |
| Sleep Disorder | 816 (23.07)% | 965 (27.28)% | 619 (25.2)% | 171 (15.3)% | 138 (26.44)% |
| Apathy | 1 (0.03)% | 0 (0.0)% | 1 (0.04)% | 0 (0.0)% | 0 (0.0)% |
| Hypothyroidism | 501 (14.16)% | 528 (14.93)% | 352 (14.33)% | 132 (11.81)% | 96 (18.39)% |
| Hyperthyroidism | 45 (1.27)% | 42 (1.19)% | 35 (1.43)% | 9 (0.81)% | 7 (1.34)% |
| Seizures | 17 (0.48)% | 21 (0.59)% | 12 (0.49)% | 2 (0.18)% | 3 (0.57)% |

| | | | | | |
|---|---|---|---|---|---|
| PTSD | 11 (0.31)% | 19 (0.54)% | 9 (0.37)% | 3 (0.27)% | 2 (0.38)% |
| Bipolar disorder | 19 (0.54)% | 25 (0.71)% | 16 (0.65)% | 1 (0.09)% | 1 (0.19)% |
| Schizophrenia | 4 (0.11)% | 5 (0.14)% | 4 (0.16)% | 0 (0.0)% | 0 (0.0)% |
| OCD | 7 (0.2)% | 10 (0.28)% | 4 (0.16)% | 2 (0.18)% | 3 (0.57)% |
| Arthritis | 43 (1.22)% | 54 (1.53)% | 33 (1.34)% | 8 (0.72)% | 4 (0.77)% |
| Asthma | 257 (7.27)% | 285 (8.06)% | 197 (8.02)% | 47 (4.2)% | 49 (9.39)% |
| Hearing impairment | 174 (4.92)% | 218 (6.16)% | 137 (5.58)% | 24 (2.15)% | 35 (6.7)% |
| Vision impairment | 6 (0.17)% | 2 (0.06)% | 4 (0.16)% | 2 (0.18)% | 0 (0.0)% |
| Significant alcohol use | 74 (2.09)% | 75 (2.12)% | 53 (2.16)% | 24 (2.15)% | 11 (2.11)% |
| CCI score (mean) | 1.09 | 1.14 | 1.15 | 1.06 | 1.06 |
| **Lab** | | | | | |
| Hemoglobin_10~20 | 259 (7.32)% | 186 (5.26)% | 182 (7.41)% | 93 (8.32)% | 32 (6.13)% |
| Hemoglobin_4~6.5 | 743 (21.01)% | 876 (24.77)% | 567 (23.09)% | 154 (13.77)% | 130 (24.9)% |
| Hemoglobin_6.5~8 | 575 (16.26)% | 509 (14.39)% | 429 (17.47)% | 161 (14.4)% | 106 (20.31)% |
| Hemoglobin_8~10 | 457 (12.92)% | 391 (11.05)% | 334 (13.6)% | 166 (14.85)% | 57 (10.92)% |
| Hemoglobin_<4 or >20(miss) | 1503 (42.49)% | 1575 (44.53)% | 944 (38.44)% | 544 (48.66)% | 197 (37.74)% |
| Total cholesterol_150~500 | 1411 (39.89)% | 1435 (40.57)% | 1023 (41.65)% | 388 (34.7)% | 239 (45.79)% |
| Total cholesterol_20~150 | 412 (11.65)% | 417 (11.79)% | 309 (12.58)% | 122 (10.91)% | 54 (10.34)% |
| Total cholesterol_500~880 | 0 (0.0)% | 2 (0.06)% | 0 (0.0)% | 0 (0.0)% | 0 (0.0)% |
| Total cholesterol_880~1500 | 0 (0.0)% | 0 (0.0)% | 0 (0.0)% | 0 (0.0)% | 0 (0.0)% |
| Total cholesterol_<20 or >1500(miss) | 1714 (48.46)% | 1683 (47.58)% | 1124 (45.77)% | 608 (54.38)% | 229 (43.87)% |
| LDL_100~130 | 524 (14.81)% | 502 (14.19)% | 372 (15.15)% | 136 (12.16)% | 100 (19.16)% |
| LDL_130~160 | 293 (8.28)% | 331 (9.36)% | 222 (9.04)% | 71 (6.35)% | 41 (7.85)% |
| LDL_160~190 | 115 (3.25)% | 116 (3.28)% | 81 (3.3)% | 34 (3.04)% | 14 (2.68)% |
| LDL_190~300 | 34 (0.96)% | 33 (0.93)% | 22 (0.9)% | 15 (1.34)% | 4 (0.77)% |
| LDL_20~100 | 812 (22.96)% | 799 (22.59)% | 608 (24.76)% | 237 (21.2)% | 129 (24.71)% |
| LDL_<20 or >300(miss) | 1759 (49.73)% | 1756 (49.65)% | 1151 (46.86)% | 625 (55.9)% | 234 (44.83)% |
| eGFR_<15 | 3 (0.08)% | 4 (0.11)% | 1 (0.04)% | 2 (0.18)% | 0 (0.0)% |
| eGFR_15~30 | 2 (0.06)% | 0 (0.0)% | 0 (0.0)% | 1 (0.09)% | 0 (0.0)% |
| eGFR_30~60 | 75 (2.12)% | 81 (2.29)% | 50 (2.04)% | 32 (2.86)% | 7 (1.34)% |
| eGFR_60~90 | 559 (15.8)% | 592 (16.74)% | 390 (15.88)% | 195 (17.44)% | 59 (11.3)% |
| eGFR_>90 | 823 (23.27)% | 694 (19.62)% | 560 (22.8)% | 319 (28.53)% | 121 (23.18)% |
| **Medications** | | | | | |
| Phentermine/Topiramate | 23 (0.65)% | 23 (0.65)% | 14 (0.57)% | 9 (0.81)% | 3 (0.57)% |
| Bupropion/Naltrexone | 8 (0.23)% | 12 (0.34)% | 5 (0.2)% | 3 (0.27)% | 3 (0.57)% |
| Lorcaserin | 9 (0.25)% | 7 (0.2)% | 2 (0.08)% | 8 (0.72)% | 0 (0.0)% |
| Anorectic drugs | 9 (0.25)% | 7 (0.2)% | 2 (0.08)% | 8 (0.72)% | 0 (0.0)% |
| SGLT2 | 426 (12.04)% | 408 (11.54)% | 298 (12.13)% | 147 (13.15)% | 51 (9.77)% |
| Sulfonylurea | 375 (10.6)% | 369 (10.43)% | 229 (9.32)% | 167 (14.94)% | 41 (7.85)% |
| CCB | 1 (0.03)% | 3 (0.08)% | 0 (0.0)% | 1 (0.09)% | 0 (0.0)% |
| Beta blockers | 6 (0.17)% | 5 (0.14)% | 1 (0.04)% | 5 (0.45)% | 0 (0.0)% |
| Statin | 1155 (32.65)% | 1188 (33.59)% | 795 (32.37)% | 413 (36.94)% | 141 (27.01)% |
| Insulin | 534 (15.1)% | 491 (13.88)% | 317 (12.91)% | 263 (23.52)% | 61 (11.69)% |
| ACEIs | 634 (17.92)% | 625 (17.67)% | 421 (17.14)% | 263 (23.52)% | 81 (15.52)% |
| ARBs | 535 (15.13)% | 595 (16.82)% | 375 (15.27)% | 156 (13.95)% | 76 (14.56)% |
| Antidepressants | 247 (6.98)% | 251 (7.1)% | 154 (6.27)% | 82 (7.33)% | 40 (7.66)% |
| diuretic | 42 (1.19)% | 32 (0.9)% | 22 (0.9)% | 22 (1.97)% | 4 (0.77)% |
| Metformin | 1301 (36.78)% | 1271 (35.93)% | 866 (35.26)% | 542 (48.48)% | 161 (30.84)% |
| Thiazolidinedione | 97 (2.74)% | 91 (2.57)% | 66 (2.69)% | 42 (3.76)% | 13 (2.49)% |
| DPP4i | 261 (7.38)% | 266 (7.52)% | 168 (6.84)% | 110 (9.84)% | 29 (5.56)% |
| NSAIDS | 480 (13.57)% | 562 (15.89)% | 339 (13.8)% | 136 (12.16)% | 74 (14.18)% |
| Antipsychotics | 33 (0.93)% | 46 (1.3)% | 17 (0.69)% | 14 (1.25)% | 6 (1.15)% |
| Alpha glucosidase inhibitor | 4 (0.11)% | 2 (0.06)% | 3 (0.12)% | 2 (0.18)% | 0 (0.0)% |
| Meglitinide | 9 (0.25)% | 5 (0.14)% | 7 (0.29)% | 2 (0.18)% | 1 (0.19)% |
| Anti-dementia | 5 (0.14)% | 12 (0.34)% | 2 (0.08)% | 1 (0.09)% | 1 (0.19)% |
| Anticoagulant | 38 (1.07)% | 37 (1.05)% | 25 (1.02)% | 12 (1.07)% | 3 (0.57)% |
| Antiplatelets | 187 (5.29)% | 188 (5.32)% | 113 (4.6)% | 95 (8.5)% | 16 (3.07)% |
| Aldosterone receptor antagonists | 38 (1.07)% | 32 (0.9)% | 21 (0.86)% | 19 (1.7)% | 4 (0.77)% |
| Opioids | 273 (7.72)% | 309 (8.74)% | 169 (6.88)% | 102 (9.12)% | 38 (7.28)% |

| | | | | | |
|---|---|---|---|---|---|
| Oral corticosteroids | 645 (18.24)% | 706 (19.96)% | 452 (18.4)% | 180 (16.1)% | 95 (18.2)% |
| Orlistat | 1 (0.03)% | 1 (0.03)% | 1 (0.04)% | 0 (0.0)% | 0 (0.0)% |

GLP-1RAs, glucagon-like peptide-1 receptor agonists; DPP4i, dipeptidyl peptidase 4 inhibitors; SGLT2 Inhibitors, sodium-glucose cotransporter 2 inhibitors; ACEI, angiotensin-converting-enzyme inhibitors; ARB, angiotensin II receptor blockers; NSAIDS, non-steroidal anti-inflammatory drugs; CCB, Calcium channel blockers; GERD, gastroesophageal reflux disease; NAFLD, nonalcoholic fatty liver disease; NASH, nonalcoholic steatohepatitis; OCD, Obsessive-compulsive disorder; PTSD, Post-traumatic stress disorder; CCI, Carlson comorbidity index; Others: include American Indian or Alaska Native, Asian, Native Hawaiian or Other Pacific Islander, and individuals identifying as multiracial.

**eTable 7. GLP1-RAs treatment effect baseline characteristics in subphenotype 2**

| Variable | Users, N (%) | Non-Users, N (%) | SEMA_user, N (%) | LIRA_user, N (%) | TIRZ_user, N (%) |
|---|---|---|---|---|---|
| Total | 1909 (50.0)% | 1909 (50.0)% | 1189 (62.28)% | 763 (39.97)% | 242 (12.68)% |
| AGE (mean) | 51.51 | 51.72 | 51.91 | 51.13 | 49.71 |
| **Vital** | | | | | |
| BMI (mean) | 35.81 | 36.4 | 35.96 | 35.84 | 35.63 |
| Systolic blood pressure (mean) | 130.95 | 130.37 | 131.35 | 130.71 | 130.74 |
| Diastolic blood pressure (mean) | 80 | 80.17 | 80.04 | 79.73 | 80.74 |
| **Race Ethnicity** | | | | | |
| Hispanic | 203 (10.63)% | 190 (9.95)% | 141 (11.86)% | 65 (8.52)% | 41 (16.94)% |
| NHB | 805 (42.17)% | 859 (45.0)% | 512 (43.06)% | 311 (40.76)% | 88 (36.36)% |
| NHW | 769 (40.28)% | 738 (38.66)% | 442 (37.17)% | 350 (45.87)% | 94 (38.84)% |
| Others | 59 (3.09)% | 57 (2.99)% | 44 (3.7)% | 20 (2.62)% | 4 (1.65)% |
| Unknown | 73 (3.82)% | 65 (3.4)% | 50 (4.21)% | 17 (2.23)% | 15 (6.2)% |
| **Sex** | | | | | |
| Female | 1546 (80.98)% | 1551 (81.25)% | 967 (81.33)% | 606 (79.42)% | 203 (83.88)% |
| Male | 363 (19.02)% | 358 (18.75)% | 222 (18.67)% | 157 (20.58)% | 39 (16.12)% |
| **Comorbidities** | | | | | |
| Pulmonary Embolism | 28 (1.47)% | 34 (1.78)% | 22 (1.85)% | 7 (0.92)% | 0 (0.0)% |
| atrial fibrillation CCW | 40 (2.1)% | 56 (2.93)% | 27 (2.27)% | 14 (1.83)% | 5 (2.07)% |
| Venous Thromboembolism | 47 (2.46)% | 56 (2.93)% | 34 (2.86)% | 13 (1.7)% | 3 (1.24)% |
| Hypertension | 1272 (66.63)% | 1287 (67.42)% | 834 (70.14)% | 455 (59.63)% | 162 (66.94)% |
| Hyperlipidemia CCW | 1174 (61.5)% | 1175 (61.55)% | 806 (67.79)% | 383 (50.2)% | 158 (65.29)% |
| Type 2 Diabetes CCW | 1076 (56.36)% | 1031 (54.01)% | 706 (59.38)% | 424 (55.57)% | 124 (51.24)% |
| GERD | 824 (43.16)% | 875 (45.84)% | 559 (47.01)% | 251 (32.9)% | 113 (46.69)% |
| Depression | 700 (36.67)% | 735 (38.5)% | 477 (40.12)% | 205 (26.87)% | 108 (44.63)% |
| Osteoarthritis | 634 (33.21)% | 737 (38.61)% | 449 (37.76)% | 173 (22.67)% | 82 (33.88)% |
| Vitamin D deficiency | 695 (36.41)% | 755 (39.55)% | 467 (39.28)% | 216 (28.31)% | 114 (47.11)% |
| Sleep apnea | 505 (26.45)% | 542 (28.39)% | 356 (29.94)% | 134 (17.56)% | 85 (35.12)% |
| prediabetes | 712 (37.3)% | 798 (41.8)% | 497 (41.8)% | 191 (25.03)% | 115 (47.52)% |
| Gallbladder Disease | 18 (0.94)% | 22 (1.15)% | 13 (1.09)% | 8 (1.05)% | 1 (0.41)% |
| Metabolic Syndrome | 60 (3.14)% | 69 (3.61)% | 41 (3.45)% | 18 (2.36)% | 13 (5.37)% |
| Dyspepsia | 31 (1.62)% | 27 (1.41)% | 19 (1.6)% | 11 (1.44)% | 5 (2.07)% |
| Inflammatory Bowel Disease | 23 (1.2)% | 21 (1.1)% | 16 (1.35)% | 8 (1.05)% | 4 (1.65)% |
| Acute/Chronic Pancreatitis | 14 (0.73)% | 16 (0.84)% | 10 (0.84)% | 3 (0.39)% | 3 (1.24)% |
| Anorexia | 22 (1.15)% | 31 (1.62)% | 19 (1.6)% | 2 (0.26)% | 5 (2.07)% |
| Cushing Syndrome | 8 (0.42)% | 5 (0.26)% | 2 (0.17)% | 3 (0.39)% | 3 (1.24)% |
| Feeding Difficulties | 351 (18.39)% | 375 (19.64)% | 274 (23.04)% | 69 (9.04)% | 67 (27.69)% |
| Prader-Willi Syndrome | 0 (0.0)% | 1 (0.05)% | 0 (0.0)% | 0 (0.0)% | 0 (0.0)% |
| Congestive heart failure | 9 (0.47)% | 10 (0.52)% | 5 (0.42)% | 3 (0.39)% | 2 (0.83)% |
| Peripheral vascular disease | 93 (4.87)% | 117 (6.13)% | 62 (5.21)% | 32 (4.19)% | 9 (3.72)% |
| Cerebrovascular disease | 33 (1.73)% | 43 (2.25)% | 23 (1.93)% | 8 (1.05)% | 2 (0.83)% |
| Dementia | 3 (0.16)% | 5 (0.26)% | 2 (0.17)% | 1 (0.13)% | 0 (0.0)% |
| Chronic pulmonary disease | 526 (27.55)% | 573 (30.02)% | 356 (29.94)% | 162 (21.23)% | 74 (30.58)% |
| Rheumatic disease | 105 (5.5)% | 124 (6.5)% | 64 (5.38)% | 37 (4.85)% | 15 (6.2)% |
| Peptic ulcer disease | 47 (2.46)% | 69 (3.61)% | 31 (2.61)% | 15 (1.97)% | 8 (3.31)% |
| Mild liver disease | 46 (2.41)% | 57 (2.99)% | 30 (2.52)% | 13 (1.7)% | 3 (1.24)% |
| Diabetes without chronic complication | 1069 (56.0)% | 1037 (54.32)% | 707 (59.46)% | 418 (54.78)% | 123 (50.83)% |
| Diabetes with chronic complication | 366 (19.17)% | 367 (19.22)% | 247 (20.77)% | 125 (16.38)% | 37 (15.29)% |
| Hemiplegia or paraplegia | 13 (0.68)% | 18 (0.94)% | 8 (0.67)% | 1 (0.13)% | 5 (2.07)% |
| Renal disease | 3 (0.16)% | 6 (0.31)% | 1 (0.08)% | 1 (0.13)% | 1 (0.41)% |
| AIDS/HIV | 31 (1.62)% | 30 (1.57)% | 13 (1.09)% | 17 (2.23)% | 3 (1.24)% |
| Deep Vein Thrombosis (DVT) | 19 (1.0)% | 21 (1.1)% | 12 (1.01)% | 6 (0.79)% | 3 (1.24)% |
| Anxiety | 819 (42.9)% | 895 (46.88)% | 578 (48.61)% | 222 (29.1)% | 125 (51.65)% |
| Sleep Disorder | 859 (45.0)% | 944 (49.45)% | 593 (49.87)% | 248 (32.5)% | 135 (55.79)% |
| Apathy | 1 (0.05)% | 0 (0.0)% | 1 (0.08)% | 0 (0.0)% | 0 (0.0)% |
| Hypothyroidism | 343 (17.97)% | 356 (18.65)% | 241 (20.27)% | 104 (13.63)% | 50 (20.66)% |
| Hyperthyroidism | 46 (2.41)% | 49 (2.57)% | 35 (2.94)% | 15 (1.97)% | 7 (2.89)% |

| | | | | | |
|---|---|---|---|---|---|
| Seizures | 28 (1.47)% | 38 (1.99)% | 16 (1.35)% | 10 (1.31)% | 6 (2.48)% |
| PTSD | 52 (2.72)% | 76 (3.98)% | 36 (3.03)% | 14 (1.83)% | 8 (3.31)% |
| Bipolar disorder | 65 (3.4)% | 71 (3.72)% | 54 (4.54)% | 15 (1.97)% | 7 (2.89)% |
| Schizophrenia | 25 (1.31)% | 21 (1.1)% | 20 (1.68)% | 5 (0.66)% | 4 (1.65)% |
| OCD | 19 (1.0)% | 22 (1.15)% | 14 (1.18)% | 6 (0.79)% | 4 (1.65)% |
| Arthritis | 71 (3.72)% | 82 (4.3)% | 42 (3.53)% | 24 (3.15)% | 9 (3.72)% |
| Asthma | 429 (22.47)% | 472 (24.72)% | 298 (25.06)% | 124 (16.25)% | 63 (26.03)% |
| Hearing impairment | 119 (6.23)% | 166 (8.7)% | 80 (6.73)% | 30 (3.93)% | 21 (8.68)% |
| Vision impairment | 10 (0.52)% | 10 (0.52)% | 7 (0.59)% | 3 (0.39)% | 1 (0.41)% |
| Significant alcohol use | 103 (5.4)% | 84 (4.4)% | 61 (5.13)% | 49 (6.42)% | 4 (1.65)% |
| CCI score (mean) | 1.51 | 1.57 | 1.57 | 1.37 | 1.4 |
| **Lab** | | | | | |
| Hemoglobin_10~20 | 163 (8.54)% | 126 (6.6)% | 104 (8.75)% | 68 (8.91)% | 19 (7.85)% |
| Hemoglobin_4~6.5 | 509 (26.66)% | 542 (28.39)% | 332 (27.92)% | 160 (20.97)% | 83 (34.3)% |
| Hemoglobin_6.5~8 | 293 (15.35)% | 303 (15.87)% | 203 (17.07)% | 97 (12.71)% | 29 (11.98)% |
| Hemoglobin_8~10 | 220 (11.52)% | 184 (9.64)% | 131 (11.02)% | 110 (14.42)% | 28 (11.57)% |
| Hemoglobin_<4 or >20(miss) | 724 (37.93)% | 754 (39.5)% | 419 (35.24)% | 328 (42.99)% | 83 (34.3)% |
| Total cholesterol_150~500 | 810 (42.43)% | 843 (44.16)% | 513 (43.15)% | 303 (39.71)% | 116 (47.93)% |
| Total cholesterol_20~150 | 230 (12.05)% | 238 (12.47)% | 144 (12.11)% | 93 (12.19)% | 23 (9.5)% |
| Total cholesterol_500~880 | 0 (0.0)% | 0 (0.0)% | 0 (0.0)% | 0 (0.0)% | 0 (0.0)% |
| Total cholesterol_880~1500 | 0 (0.0)% | 0 (0.0)% | 0 (0.0)% | 0 (0.0)% | 0 (0.0)% |
| Total cholesterol_<20 or >1500(miss) | 869 (45.52)% | 828 (43.37)% | 532 (44.74)% | 367 (48.1)% | 103 (42.56)% |
| LDL_100~130 | 311 (16.29)% | 320 (16.76)% | 199 (16.74)% | 120 (15.73)% | 45 (18.6)% |
| LDL_130~160 | 156 (8.17)% | 171 (8.96)% | 98 (8.24)% | 60 (7.86)% | 18 (7.44)% |
| LDL_160~190 | 65 (3.4)% | 59 (3.09)% | 33 (2.78)% | 26 (3.41)% | 14 (5.79)% |
| LDL_190~300 | 12 (0.63)% | 19 (1.0)% | 8 (0.67)% | 3 (0.39)% | 2 (0.83)% |
| LDL_20~100 | 470 (24.62)% | 484 (25.35)% | 305 (25.65)% | 170 (22.28)% | 56 (23.14)% |
| LDL_<20 or >300(miss) | 895 (46.88)% | 856 (44.84)% | 546 (45.92)% | 384 (50.33)% | 107 (44.21)% |
| eGFR_<15 | 2 (0.1)% | 4 (0.21)% | 0 (0.0)% | 1 (0.13)% | 0 (0.0)% |
| eGFR_15~30 | 0 (0.0)% | 1 (0.05)% | 0 (0.0)% | 0 (0.0)% | 0 (0.0)% |
| eGFR_30~60 | 85 (4.45)% | 88 (4.61)% | 39 (3.28)% | 51 (6.68)% | 3 (1.24)% |
| eGFR_60~90 | 399 (20.9)% | 420 (22.0)% | 242 (20.35)% | 162 (21.23)% | 35 (14.46)% |
| eGFR_>90 | 538 (28.18)% | 510 (26.72)% | 322 (27.08)% | 238 (31.19)% | 74 (30.58)% |
| **Medications** | | | | | |
| Phentermine/Topiramate | 9 (0.47)% | 11 (0.58)% | 4 (0.34)% | 6 (0.79)% | 3 (1.24)% |
| Bupropion/Naltrexone | 12 (0.63)% | 11 (0.58)% | 7 (0.59)% | 6 (0.79)% | 4 (1.65)% |
| Lorcaserin | 5 (0.26)% | 4 (0.21)% | 0 (0.0)% | 5 (0.66)% | 0 (0.0)% |
| Anorectic drugs | 5 (0.26)% | 4 (0.21)% | 0 (0.0)% | 5 (0.66)% | 0 (0.0)% |
| SGLT2 | 156 (8.17)% | 139 (7.28)% | 110 (9.25)% | 57 (7.47)% | 18 (7.44)% |
| Sulfonylurea | 191 (10.01)% | 181 (9.48)% | 105 (8.83)% | 105 (13.76)% | 18 (7.44)% |
| CCB | 4 (0.21)% | 8 (0.42)% | 0 (0.0)% | 4 (0.52)% | 0 (0.0)% |
| Beta blockers | 4 (0.21)% | 5 (0.26)% | 2 (0.17)% | 2 (0.26)% | 0 (0.0)% |
| Statin | 621 (32.53)% | 576 (30.17)% | 390 (32.8)% | 261 (34.21)% | 68 (28.1)% |
| Insulin | 434 (22.73)% | 404 (21.16)% | 224 (18.84)% | 249 (32.63)% | 41 (16.94)% |
| ACEIs | 383 (20.06)% | 359 (18.81)% | 217 (18.25)% | 189 (24.77)% | 40 (16.53)% |
| ARBs | 357 (18.7)% | 340 (17.81)% | 227 (19.09)% | 146 (19.13)% | 42 (17.36)% |
| Antidepressants | 388 (20.32)% | 378 (19.8)% | 220 (18.5)% | 168 (22.02)% | 60 (24.79)% |
| diuretic | 48 (2.51)% | 68 (3.56)% | 32 (2.69)% | 22 (2.88)% | 2 (0.83)% |
| Metformin | 652 (34.15)% | 566 (29.65)% | 405 (34.06)% | 304 (39.84)% | 61 (25.21)% |
| Thiazolidinedione | 57 (2.99)% | 48 (2.51)% | 35 (2.94)% | 29 (3.8)% | 9 (3.72)% |
| DPP4i | 139 (7.28)% | 126 (6.6)% | 84 (7.06)% | 72 (9.44)% | 10 (4.13)% |
| NSAIDS | 634 (33.21)% | 635 (33.26)% | 396 (33.31)% | 256 (33.55)% | 75 (30.99)% |
| Antipsychotics | 102 (5.34)% | 91 (4.77)% | 65 (5.47)% | 47 (6.16)% | 15 (6.2)% |
| Alpha glucosidase inhibitor | 1 (0.05)% | 1 (0.05)% | 1 (0.08)% | 0 (0.0)% | 0 (0.0)% |
| Meglitinide | 3 (0.16)% | 3 (0.16)% | 1 (0.08)% | 0 (0.0)% | 1 (0.41)% |
| Anti-dementia | 3 (0.16)% | 1 (0.05)% | 2 (0.17)% | 0 (0.0)% | 1 (0.41)% |
| Anticoagulant | 44 (2.3)% | 52 (2.72)% | 22 (1.85)% | 24 (3.15)% | 5 (2.07)% |
| Antiplatelets | 200 (10.48)% | 196 (10.27)% | 111 (9.34)% | 102 (13.37)% | 23 (9.5)% |
| Aldosterone receptor antagonists | 44 (2.3)% | 58 (3.04)% | 31 (2.61)% | 19 (2.49)% | 1 (0.41)% |

| | | | | | |
|---|---|---|---|---|---|
| Opioids | 542 (28.39)% | 556 (29.13)% | 299 (25.15)% | 255 (33.42)% | 53 (21.9)% |
| Oral corticosteroids | 799 (41.85)% | 829 (43.43)% | 505 (42.47)% | 302 (39.58)% | 113 (46.69)% |
| Orlistat | 4 (0.21)% | 2 (0.1)% | 2 (0.17)% | 3 (0.39)% | 0 (0.0)% |

GLP-1RAs, glucagon-like peptide-1 receptor agonists; DPP4i, dipeptidyl peptidase 4 inhibitors; SGLT2 Inhibitors, sodium-glucose cotransporter 2 inhibitors; ACEI, angiotensin-converting-enzyme inhibitors; ARB, angiotensin II receptor blockers; NSAIDS, non-steroidal anti-inflammatory drugs; CCB, Calcium channel blockers; GERD, gastroesophageal reflux disease; NAFLD, nonalcoholic fatty liver disease; NASH, nonalcoholic steatohepatitis; OCD, Obsessive-compulsive disorder; PTSD, Post-traumatic stress disorder; CCI, Carlson comorbidity index; Others: include American Indian or Alaska Native, Asian, Native Hawaiian or Other Pacific Islander, and individuals identifying as multiracial.

**eTable 8. GLP1-RAs treatment effect baseline characteristics in subphenotype 3**

| Variable | Users, N (%) | Non-Users, N (%) | SEMA_user, N (%) | LIRA_user, N (%) | TIRZ_user, N (%) |
|---|---|---|---|---|---|
| Total | 3339 (50.0)% | 3339 (50.0)% | 2089 (62.56)% | 1172 (35.1)% | 514 (15.39)% |
| AGE (mean) | 45.12 | 46.04 | 44.78 | 45.98 | 44.08 |
| **Vital** | | | | | |
| BMI (mean) | 42.93 | 43.16 | 43.08 | 42.9 | 43.5 |
| Systolic blood pressure (mean) | 131.54 | 131.49 | 132.08 | 131.1 | 132.11 |
| Diastolic blood pressure (mean) | 81.33 | 81.53 | 81.84 | 80.4 | 81.93 |
| **Race Ethnicity** | | | | | |
| Hispanic | 324 (9.7)% | 256 (7.67)% | 233 (11.15)% | 77 (6.57)% | 52 (10.12)% |
| NHB | 996 (29.83)% | 1122 (33.6)% | 642 (30.73)% | 330 (28.16)% | 136 (26.46)% |
| NHW | 1773 (53.1)% | 1698 (50.85)% | 1062 (50.84)% | 674 (57.51)% | 290 (56.42)% |
| Others | 60 (1.8)% | 66 (1.98)% | 48 (2.3)% | 13 (1.11)% | 10 (1.95)% |
| Unknown | 186 (5.57)% | 197 (5.9)% | 104 (4.98)% | 78 (6.66)% | 26 (5.06)% |
| **Sex** | | | | | |
| Female | 2650 (79.37)% | 2677 (80.17)% | 1661 (79.51)% | 923 (78.75)% | 412 (80.16)% |
| Male | 689 (20.63)% | 662 (19.83)% | 428 (20.49)% | 249 (21.25)% | 102 (19.84)% |
| **Comorbidities** | | | | | |
| Pulmonary Embolism | 34 (1.02)% | 35 (1.05)% | 27 (1.29)% | 6 (0.51)% | 5 (0.97)% |
| atrial fibrillation CCW | 61 (1.83)% | 67 (2.01)% | 39 (1.87)% | 17 (1.45)% | 13 (2.53)% |
| Venous Thromboembolism | 61 (1.83)% | 66 (1.98)% | 48 (2.3)% | 11 (0.94)% | 8 (1.56)% |
| Hypertension | 1817 (54.42)% | 1939 (58.07)% | 1205 (57.68)% | 557 (47.53)% | 306 (59.53)% |
| Hyperlipidemia CCW | 1340 (40.13)% | 1437 (43.04)% | 935 (44.76)% | 384 (32.76)% | 225 (43.77)% |
| Type 2 Diabetes CCW | 1457 (43.64)% | 1412 (42.29)% | 968 (46.34)% | 504 (43.0)% | 233 (45.33)% |
| GERD | 857 (25.67)% | 1018 (30.49)% | 590 (28.24)% | 229 (19.54)% | 137 (26.65)% |
| Depression | 920 (27.55)% | 984 (29.47)% | 634 (30.35)% | 257 (21.93)% | 143 (27.82)% |
| Osteoarthritis | 582 (17.43)% | 675 (20.22)% | 378 (18.09)% | 173 (14.76)% | 98 (19.07)% |
| Vitamin D deficiency | 829 (24.83)% | 943 (28.24)% | 543 (25.99)% | 213 (18.17)% | 164 (31.91)% |
| Sleep apnea | 728 (21.8)% | 761 (22.79)% | 527 (25.23)% | 167 (14.25)% | 141 (27.43)% |
| prediabetes | 1094 (32.76)% | 1239 (37.11)% | 787 (37.67)% | 237 (20.22)% | 215 (41.83)% |
| Gallbladder Disease | 30 (0.9)% | 41 (1.23)% | 21 (1.01)% | 9 (0.77)% | 4 (0.78)% |
| Metabolic Syndrome | 131 (3.92)% | 153 (4.58)% | 92 (4.4)% | 38 (3.24)% | 21 (4.09)% |
| Dyspepsia | 14 (0.42)% | 19 (0.57)% | 8 (0.38)% | 6 (0.51)% | 0 (0.0)% |
| Inflammatory Bowel Disease | 12 (0.36)% | 17 (0.51)% | 10 (0.48)% | 2 (0.17)% | 1 (0.19)% |
| Acute/Chronic Pancreatitis | 16 (0.48)% | 22 (0.66)% | 14 (0.67)% | 3 (0.26)% | 0 (0.0)% |
| Anorexia | 38 (1.14)% | 51 (1.53)% | 29 (1.39)% | 8 (0.68)% | 9 (1.75)% |
| Cushing Syndrome | 17 (0.51)% | 18 (0.54)% | 11 (0.53)% | 4 (0.34)% | 2 (0.39)% |
| Feeding Difficulties | 486 (14.56)% | 454 (13.6)% | 373 (17.86)% | 63 (5.38)% | 116 (22.57)% |
| Prader-Willi Syndrome | 2 (0.06)% | 2 (0.06)% | 1 (0.05)% | 2 (0.17)% | 0 (0.0)% |
| Congestive heart failure | 14 (0.42)% | 23 (0.69)% | 10 (0.48)% | 3 (0.26)% | 3 (0.58)% |
| Peripheral vascular disease | 76 (2.28)% | 115 (3.44)% | 48 (2.3)% | 27 (2.3)% | 12 (2.33)% |
| Cerebrovascular disease | 34 (1.02)% | 46 (1.38)% | 19 (0.91)% | 12 (1.02)% | 5 (0.97)% |
| Dementia | 3 (0.09)% | 4 (0.12)% | 2 (0.1)% | 1 (0.09)% | 0 (0.0)% |
| Chronic pulmonary disease | 632 (18.93)% | 694 (20.78)% | 440 (21.06)% | 156 (13.31)% | 116 (22.57)% |
| Rheumatic disease | 85 (2.55)% | 96 (2.88)% | 54 (2.58)% | 27 (2.3)% | 8 (1.56)% |
| Peptic ulcer disease | 36 (1.08)% | 51 (1.53)% | 29 (1.39)% | 5 (0.43)% | 5 (0.97)% |
| Mild liver disease | 36 (1.08)% | 40 (1.2)% | 27 (1.29)% | 11 (0.94)% | 3 (0.58)% |
| Diabetes without chronic complication | 1453 (43.52)% | 1399 (41.9)% | 966 (46.24)% | 497 (42.41)% | 232 (45.14)% |
| Diabetes with chronic complication | 289 (8.66)% | 322 (9.64)% | 178 (8.52)% | 110 (9.39)% | 41 (7.98)% |
| Hemiplegia or paraplegia | 15 (0.45)% | 20 (0.6)% | 10 (0.48)% | 3 (0.26)% | 3 (0.58)% |
| Renal disease | 7 (0.21)% | 3 (0.09)% | 5 (0.24)% | 2 (0.17)% | 0 (0.0)% |
| AIDS/HIV | 32 (0.96)% | 32 (0.96)% | 20 (0.96)% | 9 (0.77)% | 5 (0.97)% |
| Deep Vein Thrombosis (DVT) | 32 (0.96)% | 33 (0.99)% | 22 (1.05)% | 7 (0.6)% | 4 (0.78)% |
| Anxiety | 1119 (33.51)% | 1241 (37.17)% | 760 (36.38)% | 282 (24.06)% | 219 (42.61)% |
| Sleep Disorder | 1148 (34.38)% | 1262 (37.8)% | 797 (38.15)% | 291 (24.83)% | 220 (42.8)% |
| Apathy | 0 (0.0)% | 1 (0.03)% | 0 (0.0)% | 0 (0.0)% | 0 (0.0)% |
| Hypothyroidism | 442 (13.24)% | 500 (14.97)% | 299 (14.31)% | 119 (10.15)% | 72 (14.01)% |
| Hyperthyroidism | 49 (1.47)% | 62 (1.86)% | 31 (1.48)% | 16 (1.37)% | 5 (0.97)% |

| | | | | | |
|---|---|---|---|---|---|
| Seizures | 48 (1.44)% | 53 (1.59)% | 35 (1.68)% | 11 (0.94)% | 10 (1.95)% |
| PTSD | 67 (2.01)% | 83 (2.49)% | 50 (2.39)% | 15 (1.28)% | 13 (2.53)% |
| Bipolar disorder | 110 (3.29)% | 147 (4.4)% | 77 (3.69)% | 27 (2.3)% | 19 (3.7)% |
| Schizophrenia | 32 (0.96)% | 45 (1.35)% | 22 (1.05)% | 9 (0.77)% | 3 (0.58)% |
| OCD | 22 (0.66)% | 25 (0.75)% | 14 (0.67)% | 7 (0.6)% | 4 (0.78)% |
| Arthritis | 51 (1.53)% | 62 (1.86)% | 32 (1.53)% | 15 (1.28)% | 5 (0.97)% |
| Asthma | 495 (14.82)% | 549 (16.44)% | 339 (16.23)% | 124 (10.58)% | 94 (18.29)% |
| Hearing impairment | 118 (3.53)% | 164 (4.91)% | 80 (3.83)% | 24 (2.05)% | 24 (4.67)% |
| Vision impairment | 15 (0.45)% | 17 (0.51)% | 12 (0.57)% | 1 (0.09)% | 2 (0.39)% |
| Significant alcohol use | 125 (3.74)% | 131 (3.92)% | 81 (3.88)% | 45 (3.84)% | 14 (2.72)% |
| CCI score (mean) | 0.95 | 1.01 | 1.01 | 0.87 | 0.98 |
| **Lab** | | | | | |
| Hemoglobin_10~20 | 193 (5.78)% | 141 (4.22)% | 123 (5.89)% | 78 (6.66)% | 29 (5.64)% |
| Hemoglobin_4~6.5 | 860 (25.76)% | 996 (29.83)% | 577 (27.62)% | 240 (20.48)% | 170 (33.07)% |
| Hemoglobin_6.5~8 | 353 (10.57)% | 277 (8.3)% | 254 (12.16)% | 100 (8.53)% | 60 (11.67)% |
| Hemoglobin_8~10 | 213 (6.38)% | 146 (4.37)% | 143 (6.85)% | 79 (6.74)% | 31 (6.03)% |
| Hemoglobin_<4 or >20(miss) | 1720 (51.51)% | 1779 (53.28)% | 992 (47.49)% | 675 (57.59)% | 224 (43.58)% |
| Total cholesterol_150~500 | 1068 (31.99)% | 1102 (33.0)% | 734 (35.14)% | 312 (26.62)% | 208 (40.47)% |
| Total cholesterol_20~150 | 264 (7.91)% | 253 (7.58)% | 183 (8.76)% | 79 (6.74)% | 42 (8.17)% |
| Total cholesterol_500~880 | 0 (0.0)% | 0 (0.0)% | 0 (0.0)% | 0 (0.0)% | 0 (0.0)% |
| Total cholesterol_880~1500 | 0 (0.0)% | 0 (0.0)% | 0 (0.0)% | 0 (0.0)% | 0 (0.0)% |
| Total cholesterol_<20 or >1500(miss) | 2007 (60.11)% | 1984 (59.42)% | 1172 (56.1)% | 781 (66.64)% | 264 (51.36)% |
| LDL_100~130 | 437 (13.09)% | 427 (12.79)% | 291 (13.93)% | 122 (10.41)% | 88 (17.12)% |
| LDL_130~160 | 255 (7.64)% | 263 (7.88)% | 180 (8.62)% | 68 (5.8)% | 43 (8.37)% |
| LDL_160~190 | 75 (2.25)% | 80 (2.4)% | 50 (2.39)% | 21 (1.79)% | 18 (3.5)% |
| LDL_190~300 | 15 (0.45)% | 18 (0.54)% | 11 (0.53)% | 4 (0.34)% | 3 (0.58)% |
| LDL_20~100 | 537 (16.08)% | 493 (14.76)% | 365 (17.47)% | 176 (15.02)% | 96 (18.68)% |
| LDL_<20 or >300(miss) | 2020 (60.5)% | 2058 (61.64)% | 1192 (57.06)% | 781 (66.64)% | 266 (51.75)% |
| eGFR_<15 | 1 (0.03)% | 2 (0.06)% | 0 (0.0)% | 1 (0.09)% | 0 (0.0)% |
| eGFR_15~30 | 2 (0.06)% | 7 (0.21)% | 1 (0.05)% | 1 (0.09)% | 0 (0.0)% |
| eGFR_30~60 | 74 (2.22)% | 80 (2.4)% | 43 (2.06)% | 36 (3.07)% | 7 (1.36)% |
| eGFR_60~90 | 428 (12.82)% | 517 (15.48)% | 241 (11.54)% | 180 (15.36)% | 47 (9.14)% |
| eGFR_>90 | 823 (24.65)% | 742 (22.22)% | 550 (26.33)% | 283 (24.15)% | 152 (29.57)% |
| **Medications** | | | | | |
| Phentermine/Topiramate | 12 (0.36)% | 9 (0.27)% | 7 (0.34)% | 8 (0.68)% | 1 (0.19)% |
| Bupropion/Naltrexone | 21 (0.63)% | 20 (0.6)% | 13 (0.62)% | 9 (0.77)% | 5 (0.97)% |
| Lorcaserin | 6 (0.18)% | 10 (0.3)% | 3 (0.14)% | 6 (0.51)% | 1 (0.19)% |
| Anorectic drugs | 6 (0.18)% | 10 (0.3)% | 3 (0.14)% | 6 (0.51)% | 1 (0.19)% |
| SGLT2 | 196 (5.87)% | 196 (5.87)% | 117 (5.6)% | 69 (5.89)% | 30 (5.84)% |
| Sulfonylurea | 184 (5.51)% | 187 (5.6)% | 104 (4.98)% | 88 (7.51)% | 30 (5.84)% |
| CCB | 5 (0.15)% | 9 (0.27)% | 3 (0.14)% | 2 (0.17)% | 0 (0.0)% |
| Beta blockers | 11 (0.33)% | 19 (0.57)% | 4 (0.19)% | 8 (0.68)% | 0 (0.0)% |
| Statin | 610 (18.27)% | 617 (18.48)% | 385 (18.43)% | 253 (21.59)% | 77 (14.98)% |
| Insulin | 478 (14.32)% | 456 (13.66)% | 264 (12.64)% | 234 (19.97)% | 59 (11.48)% |
| ACEIs | 519 (15.54)% | 465 (13.93)% | 306 (14.65)% | 230 (19.62)% | 75 (14.59)% |
| ARBs | 395 (11.83)% | 429 (12.85)% | 266 (12.73)% | 131 (11.18)% | 69 (13.42)% |
| Antidepressants | 409 (12.25)% | 428 (12.82)% | 255 (12.21)% | 153 (13.05)% | 69 (13.42)% |
| diuretic | 97 (2.91)% | 120 (3.59)% | 62 (2.97)% | 39 (3.33)% | 14 (2.72)% |
| Metformin | 930 (27.85)% | 882 (26.42)% | 580 (27.76)% | 381 (32.51)% | 138 (26.85)% |
| Thiazolidinedione | 61 (1.83)% | 55 (1.65)% | 37 (1.77)% | 32 (2.73)% | 5 (0.97)% |
| DPP4i | 116 (3.47)% | 135 (4.04)% | 59 (2.82)% | 56 (4.78)% | 10 (1.95)% |
| NSAIDS | 664 (19.89)% | 691 (20.69)% | 429 (20.54)% | 220 (18.77)% | 115 (22.37)% |
| Antipsychotics | 137 (4.1)% | 142 (4.25)% | 91 (4.36)% | 51 (4.35)% | 16 (3.11)% |
| Alpha glucosidase inhibitor | 0 (0.0)% | 0 (0.0)% | 0 (0.0)% | 0 (0.0)% | 0 (0.0)% |
| Meglitinide | 4 (0.12)% | 2 (0.06)% | 2 (0.1)% | 4 (0.34)% | 0 (0.0)% |
| Anti-dementia | 8 (0.24)% | 17 (0.51)% | 3 (0.14)% | 5 (0.43)% | 0 (0.0)% |
| Anticoagulant | 67 (2.01)% | 75 (2.25)% | 41 (1.96)% | 21 (1.79)% | 12 (2.33)% |
| Antiplatelets | 190 (5.69)% | 211 (6.32)% | 118 (5.65)% | 91 (7.76)% | 18 (3.5)% |
| Aldosterone receptor antagonists | 93 (2.79)% | 110 (3.29)% | 62 (2.97)% | 35 (2.99)% | 14 (2.72)% |

| | | | | | |
|---|---|---|---|---|---|
| Opioids | 525 (15.72)% | 509 (15.24)% | 293 (14.03)% | 225 (19.2)% | 70 (13.62)% |
| Oral corticosteroids | 888 (26.59)% | 961 (28.78)% | 579 (27.72)% | 271 (23.12)% | 153 (29.77)% |
| Orlistat | 3 (0.09)% | 1 (0.03)% | 3 (0.14)% | 1 (0.09)% | 0 (0.0)% |

GLP-1RAs, glucagon-like peptide-1 receptor agonists; DPP4i, dipeptidyl peptidase 4 inhibitors; SGLT2 Inhibitors, sodium-glucose cotransporter 2 inhibitors; ACEI, angiotensin-converting-enzyme inhibitors; ARB, angiotensin II receptor blockers; NSAIDS, non-steroidal anti-inflammatory drugs; CCB, Calcium channel blockers; GERD, gastroesophageal reflux disease; NAFLD, nonalcoholic fatty liver disease; NASH, nonalcoholic steatohepatitis; OCD, Obsessive-compulsive disorder; PTSD, Post-traumatic stress disorder; CCI, Carlson comorbidity index; Others: include American Indian or Alaska Native, Asian, Native Hawaiian or Other Pacific Islander, and individuals identifying as multiracial.

**eFigure 1. Davies-Bouldin Index for time series k-means clustering (K = 2 to 10) utilizing embeddings from GraphSAGE and MagNet.**

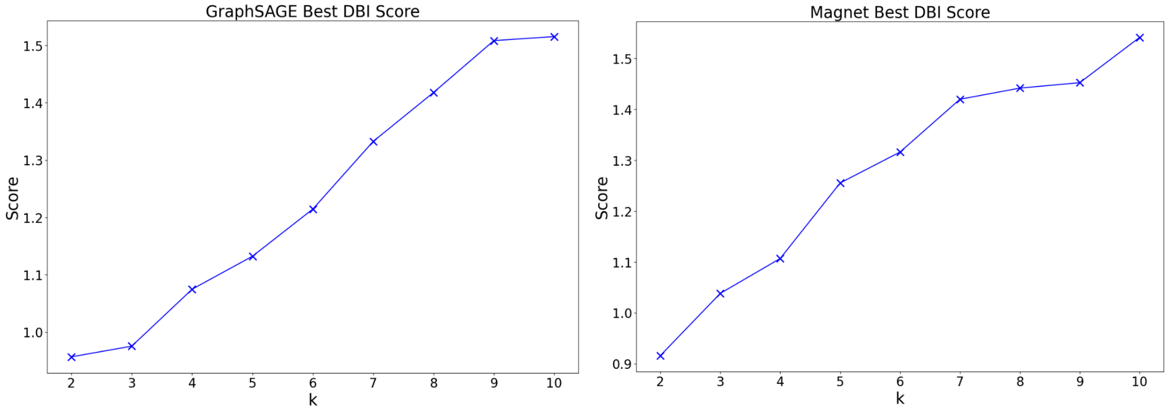

**eFigure 2. Characteristics of obesity progression subphenotypes identified by GraphSAGE with time series K-means (K = 3). (a) BTG distribution in each subphenotype. (b) Cumulative incidence of obesity related comorbidities and Kaplan-Meier survival curves for each subphenotype.**

(a)

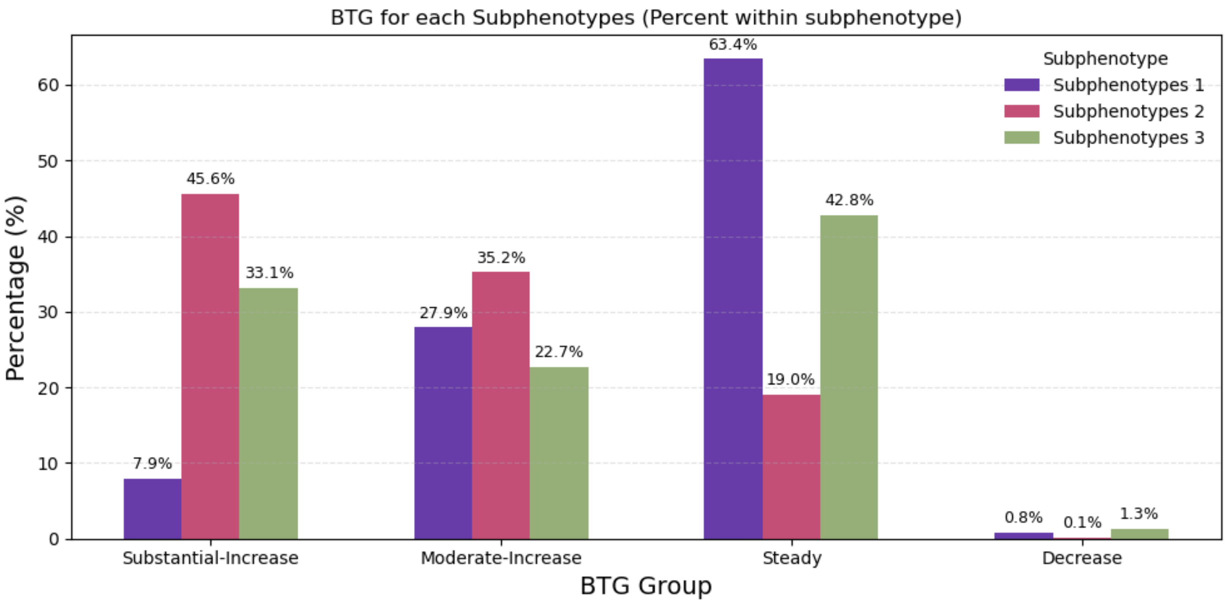


(b)

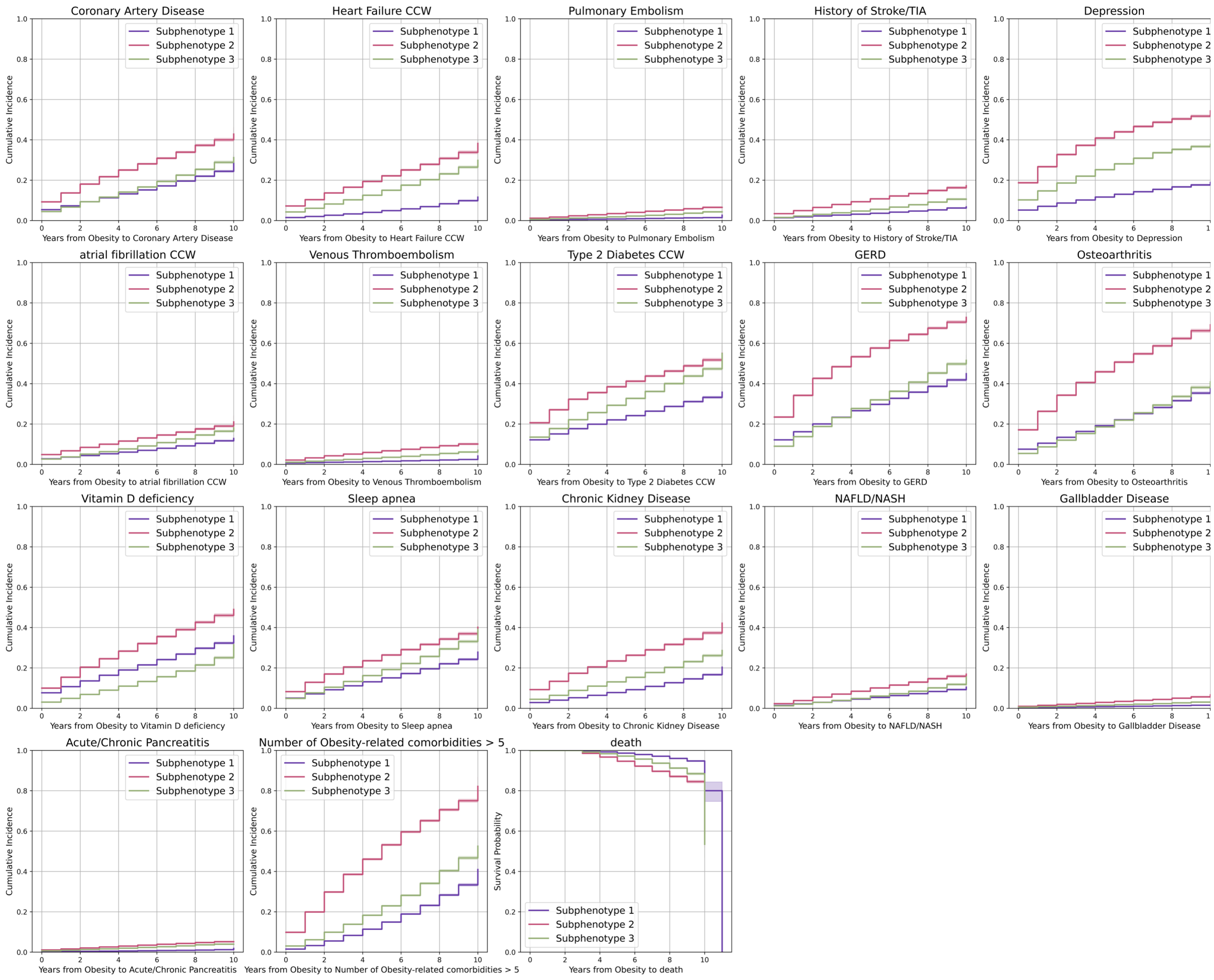

**eFigure 3. Characteristics of obesity progression subphenotypes identified by GraphSAGE with time series K-means (K = 3). (a) Demographic characteristics of each subphenotype. (b) Heatmap and circle plot of the obesity related comorbidities prevalence across the subphenotypes.**

(a)

| Subphenotype 1 (N = 102244) | | Subphenotype 2 (N = 54050) | | Subphenotype 3 (N = 80809) | |
|---|---|---|---|---|---|
| **Observation period, mean years (std)** | 4.91 (2.23) | **Observation period, mean years (std)** | 5.02 (2.29) | **Observation period, mean years (std)** | 4.85 (2.27) |
| **Age, mean (std)** | 56.45 (15.24) | **Age, mean (std)** | 52.39 (17.34) | **Age, mean (std)** | 45.48 (17.63) |
| **Female, N (%)** | 47693 (46.65%) | **Female, N (%)** | 33777 (62.49%) | **Female, N (%)** | 56220 (69.57%) |
| **Race-ethnicity, N (%)** | | **Race-ethnicity, N (%)** | | **Race-ethnicity, N (%)** | |
| Non-Hispanic White | 53636 (52.46%) | Non-Hispanic White | 23765 (43.97%) | Non-Hispanic White | 37455 (46.35%) |
| Non-Hispanic Black | 16311 (15.95%) | Non-Hispanic Black | 22045 (40.79%) | Non-Hispanic Black | 27886 (34.51%) |
| Hispanic | 17526 (17.14%) | Hispanic | 4832 (8.94%) | Hispanic | 8420 (10.42%) |
| Others | 3393 (3.32%) | Others | 1423 (2.63%) | Others | 1260 (1.56%) |
| Unknown | 11378 (11.13%) | Unknown | 1985 (3.67%) | Unknown | 5788 (7.16%) |

(b)

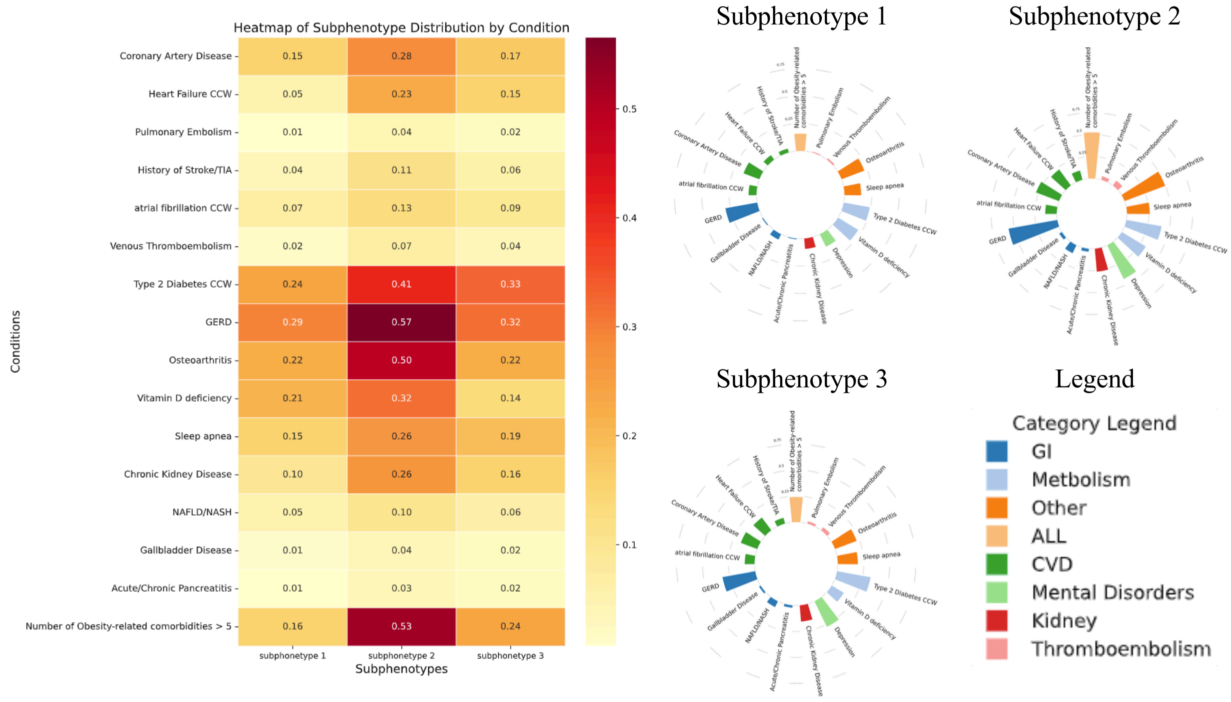

**eFigure 4. Characteristics of obesity progression subphenotypes identified by GraphSAGE with time series K-means (K = 4). (a) BTG distribution in each subphenotype. (b) Cumulative incidence of obesity related comorbidities and Kaplan-Meier survival curves for each subphenotype.**

(a)

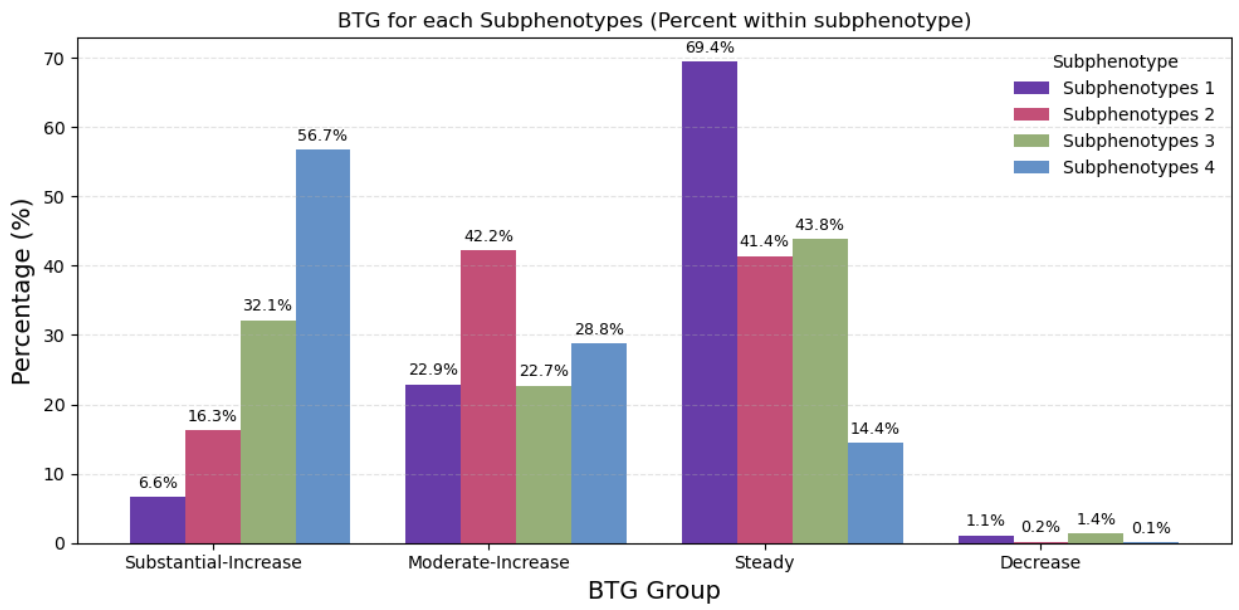


(b)

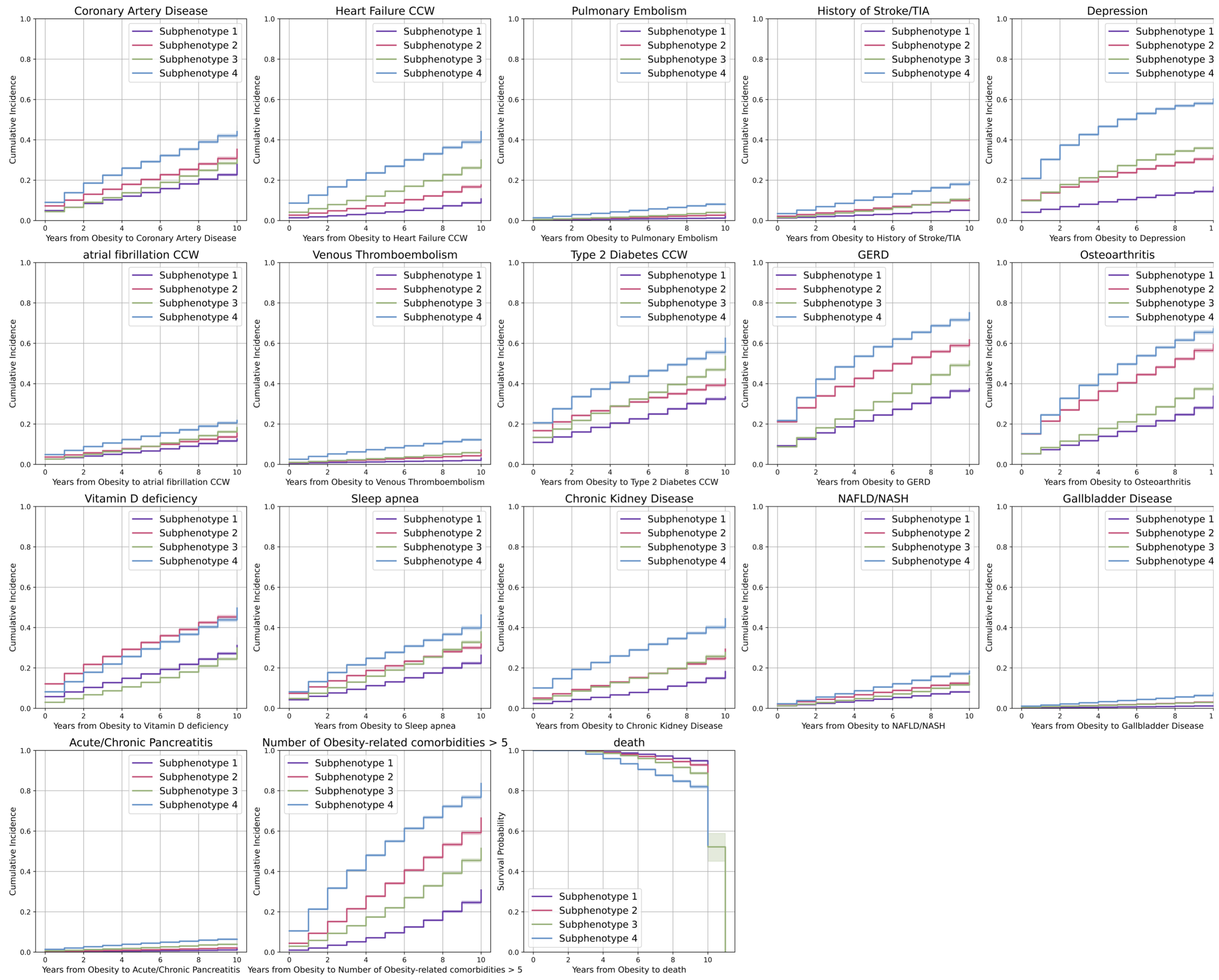

**eFigure 5. Characteristics of obesity progression subphenotypes identified by GraphSAGE with time series K-means (K = 4). (a) Demographic characteristics of each subphenotype. (b) Heatmap and circle plot of the obesity related comorbidities prevalence across the subphenotypes.**

(a)

| Subphenotype 1 (N = 74761) | | Subphenotype 2 (N = 48311) | | Subphenotype 3 (N = 72792) | | Subphenotype 4 (N = 41239) | |
|---|---|---|---|---|---|---|---|
| **Observation period, mean years (std)** | **4.84 (2.24)** | **Observation period, mean years (std)** | **5.05 (2.21)** | **Observation period, mean years (std)** | **4.85 (2.27)** | **Observation period, mean years (std)** | **5.0 (2.32)** |
| **Age, mean (std)** | **56.31 (15.3)** | **Age, mean (std)** | **56.79 (15.79)** | **Age, mean (std)** | **45.21 (17.7)** | **Age, mean (std)** | **49.32 (17.12)** |
| **Female, N (%)** | **34851 (46.62%)** | **Female, N (%)** | **24808 (51.35%)** | **Female, N (%)** | **50984 (70.04%)** | **Female, N (%)** | **27047 (65.59%)** |
| **Race-ethnicity, N (%)** | | **Race-ethnicity, N (%)** | | **Race-ethnicity, N (%)** | | **Race-ethnicity, N (%)** | |
| Non-Hispanic White | **40246 (53.83%)** | Non-Hispanic White | **23252 (48.13%)** | Non-Hispanic White | **33720 (46.32%)** | Non-Hispanic White | **17638 (42.77%)** |
| Non-Hispanic Black | **10411 (13.93%)** | Non-Hispanic Black | **12192 (25.24%)** | Non-Hispanic Black | **24993 (34.33%)** | Non-Hispanic Black | **18646 (45.21%)** |
| Hispanic | **13039 (17.44%)** | Hispanic | **7196 (14.9%)** | Hispanic | **7643 (10.5%)** | Hispanic | **2900 (7.03%)** |
| Others | **2166 (2.9%)** | Others | **1937 (4.01%)** | Others | **1125 (1.55%)** | Others | **848 (2.06%)** |
| Unknown | **8899 (11.9%)** | Unknown | **1937 (4.01%)** | Unknown | **5311 (7.3%)** | Unknown | **1207 (2.93%)** |

(b)

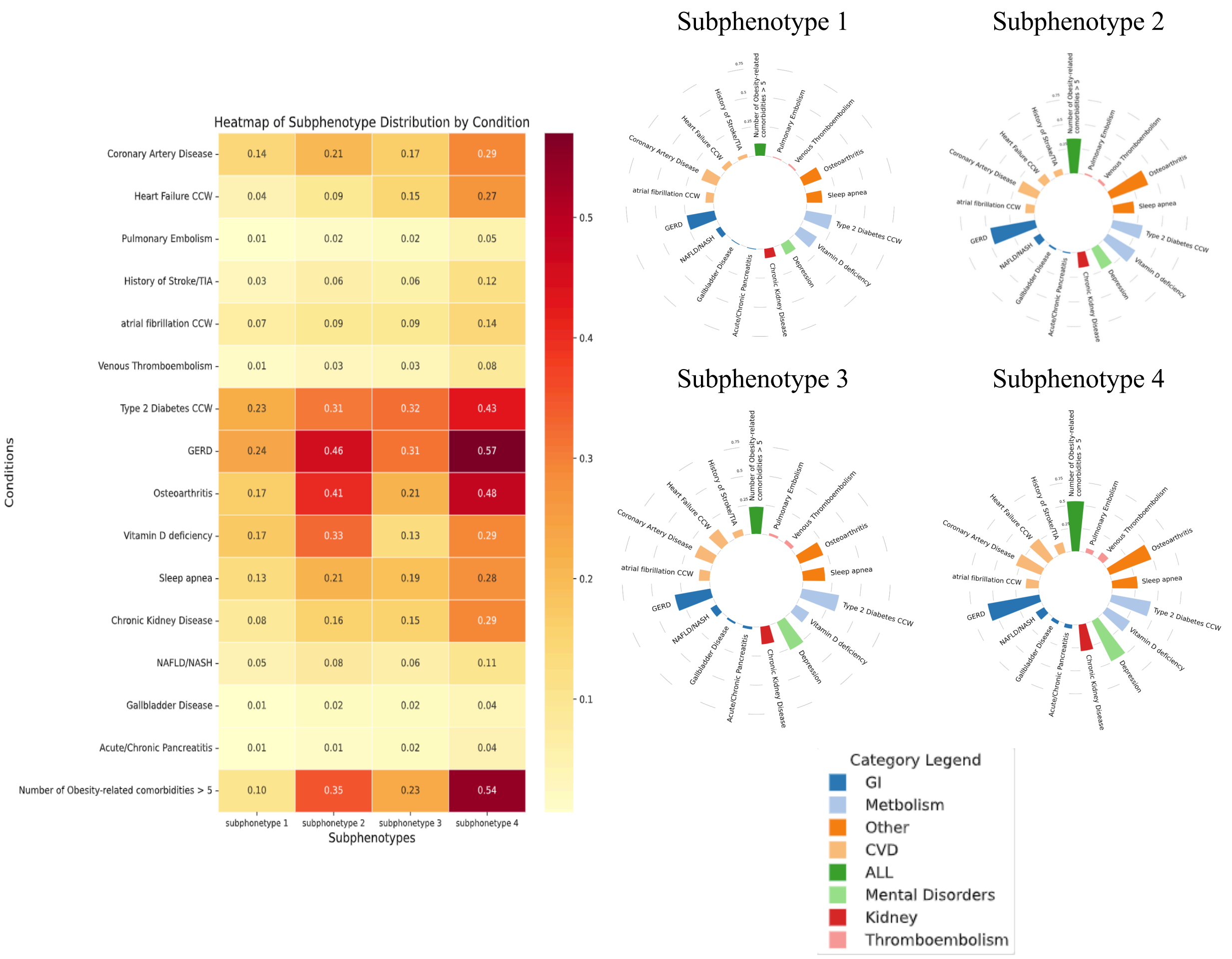

**eFigure 6. Characteristics of obesity progression subphenotypes identified by MagNet with time series K-means (K = 4). (a) BTG distribution in each subphenotype. (b) Cumulative incidence of obesity related comorbidities and Kaplan-Meier survival curves for each subphenotype.**

(a)

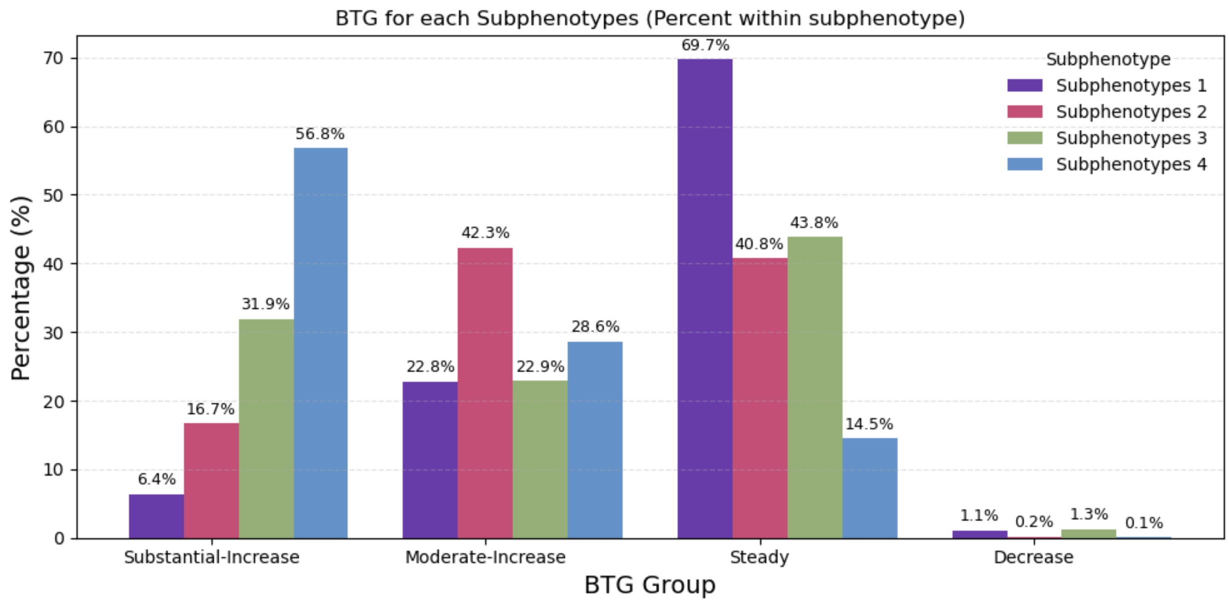


(b)

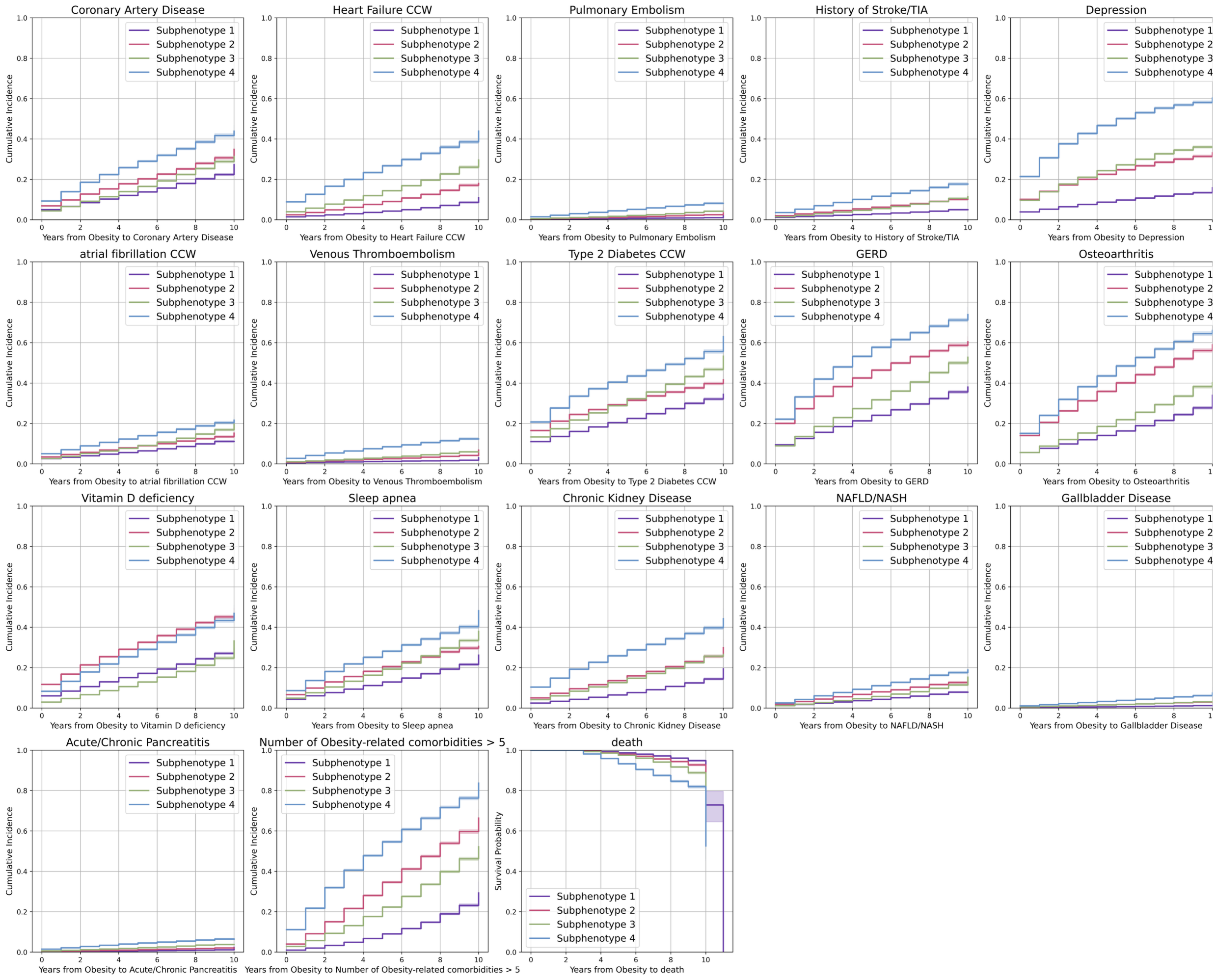

**eFigure 7. Characteristics of obesity progression subphenotypes identified by MagNet with time series K-means (K = 4). (a) Demographic characteristics of each subphenotype. (b) Heatmap and circle plot of the obesity related comorbidities prevalence across the subphenotypes.**

(a)

| Subphenotype 1 (N = 75697) | | Subphenotype 2 (N = 47768) | | Subphenotype 3 (N = 71723) | | Subphenotype 4 (N = 41915) | |
|---|---|---|---|---|---|---|---|
| **Observation period, mean years (std)** | 4.78 (2.22) | **Observation period, mean years (std)** | 5.17 (2.22) | **Observation period, mean years (std)** | 4.88 (2.27) | **Observation period, mean years (std)** | 4.93 (2.32) |
| **Age, mean (std)** | 55.96 (15.17) | **Age, mean (std)** | 56.61 (16.3) | **Age, mean (std)** | 45.7 (17.88) | **Age, mean (std)** | 49.15 (17.02) |
| **Female, N (%)** | 35344 (46.69%) | **Female, N (%)** | 25364 (53.1%) | **Female, N (%)** | 49768 (69.39%) | **Female, N (%)** | 27214 (64.93%) |
| **Race-ethnicity, N (%)** | | **Race-ethnicity, N (%)** | | **Race-ethnicity, N (%)** | | **Race-ethnicity, N (%)** | |
| Non-Hispanic White | 39747 (52.51%) | Non-Hispanic White | 22758 (47.64%) | Non-Hispanic White | 34148 (47.61%) | Non-Hispanic White | 18203 (43.43%) |
| Non-Hispanic Black | 10902 (14.4%) | Non-Hispanic Black | 12727 (26.64%) | Non-Hispanic Black | 23769 (33.14%) | Non-Hispanic Black | 18844 (44.96%) |
| Hispanic | 13532 (17.88%) | Hispanic | 6997 (14.65%) | Hispanic | 7409 (10.33%) | Hispanic | 2840 (6.78%) |
| Others | 2134 (2.82%) | Others | 1897 (3.97%) | Others | 1134 (1.58%) | Others | 911 (2.17%) |
| Unknown | 9382 (12.39%) | Unknown | 3389 (7.09%) | Unknown | 5263 (7.34%) | Unknown | 1117 (2.66%) |

(b)

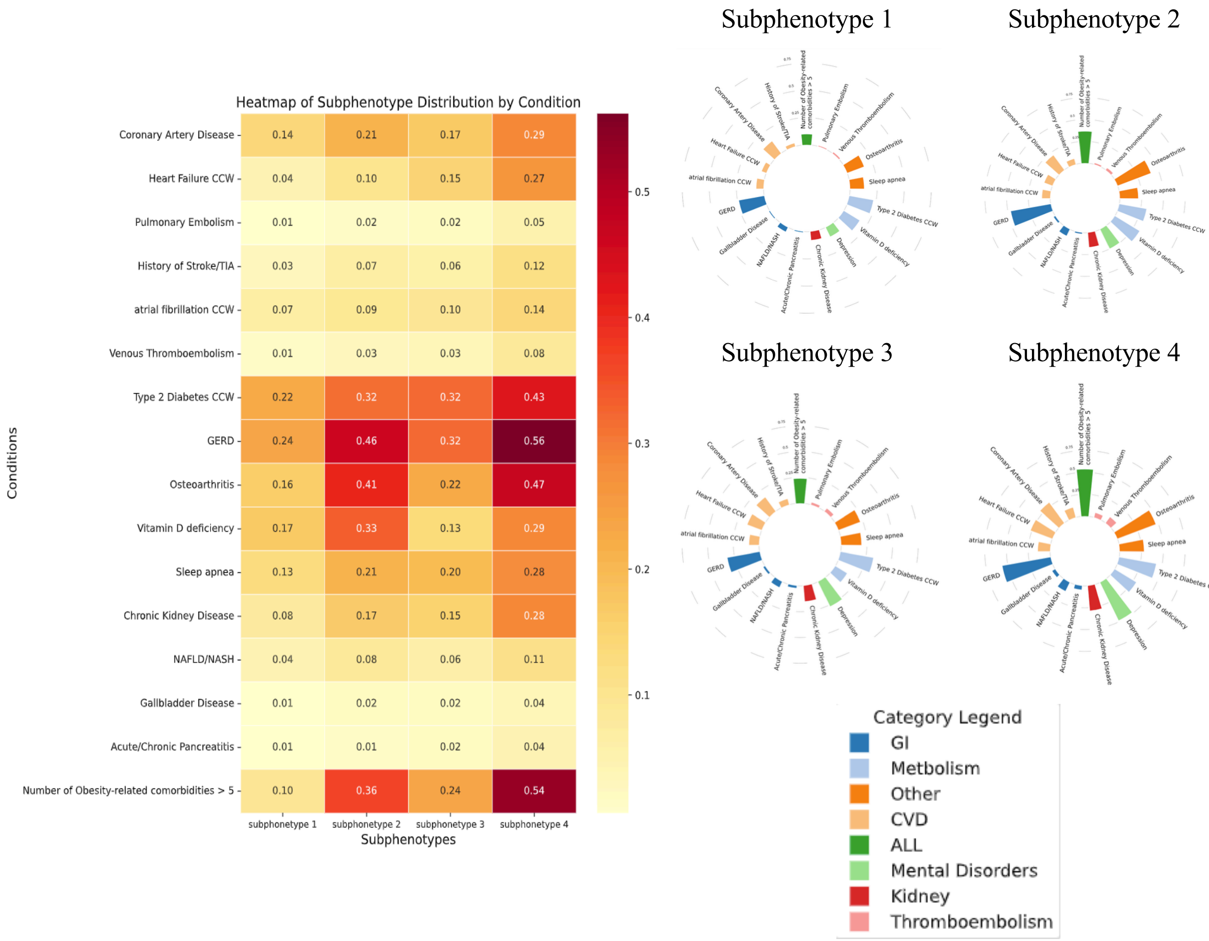

**eFigure 8. Predicting obesity subphenotypes: XGBoost vs linear models.**

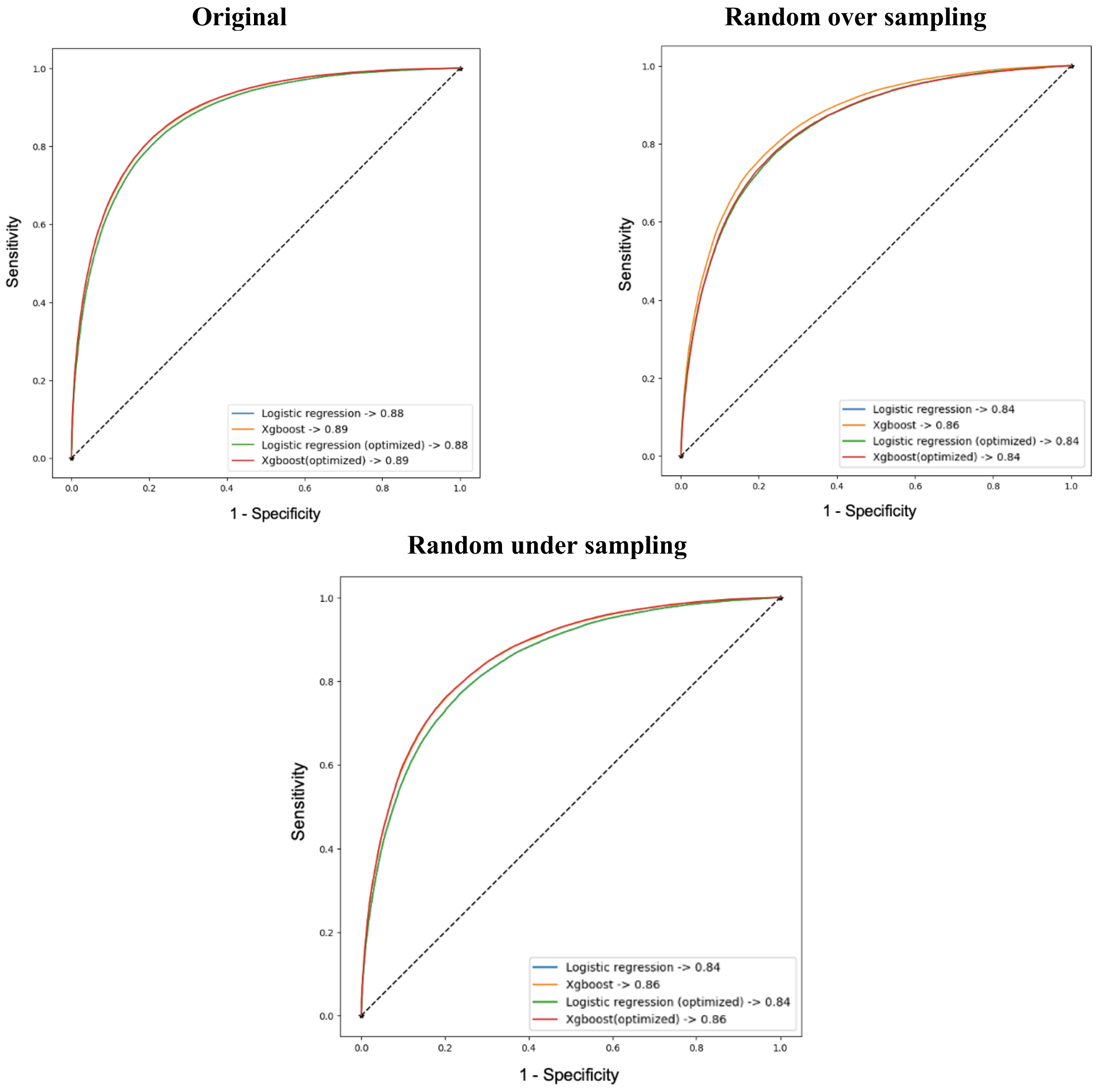


**eFigure 9. Socioeconomic status among subphenotypes in year 1.**

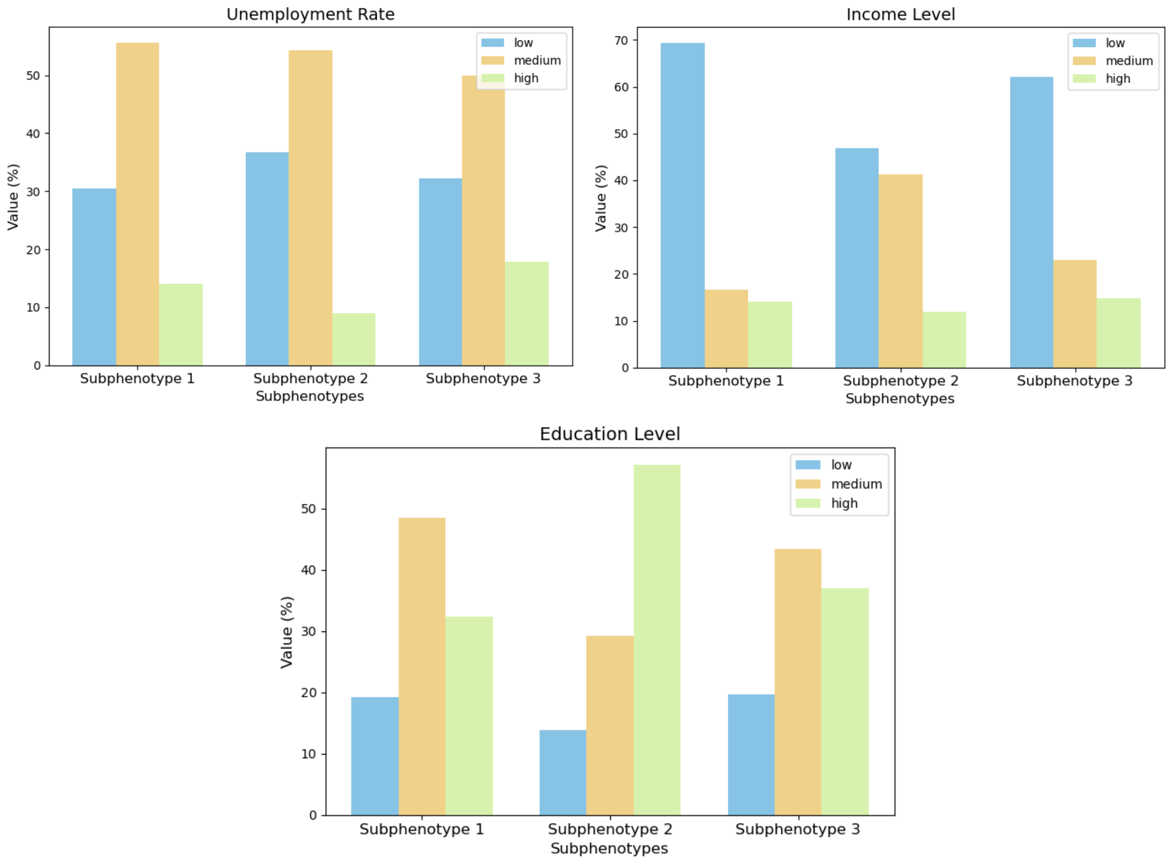

**eFigure 10. Socioeconomic status over follow-up period in (a) S1; (b) S2; (c) S3.**

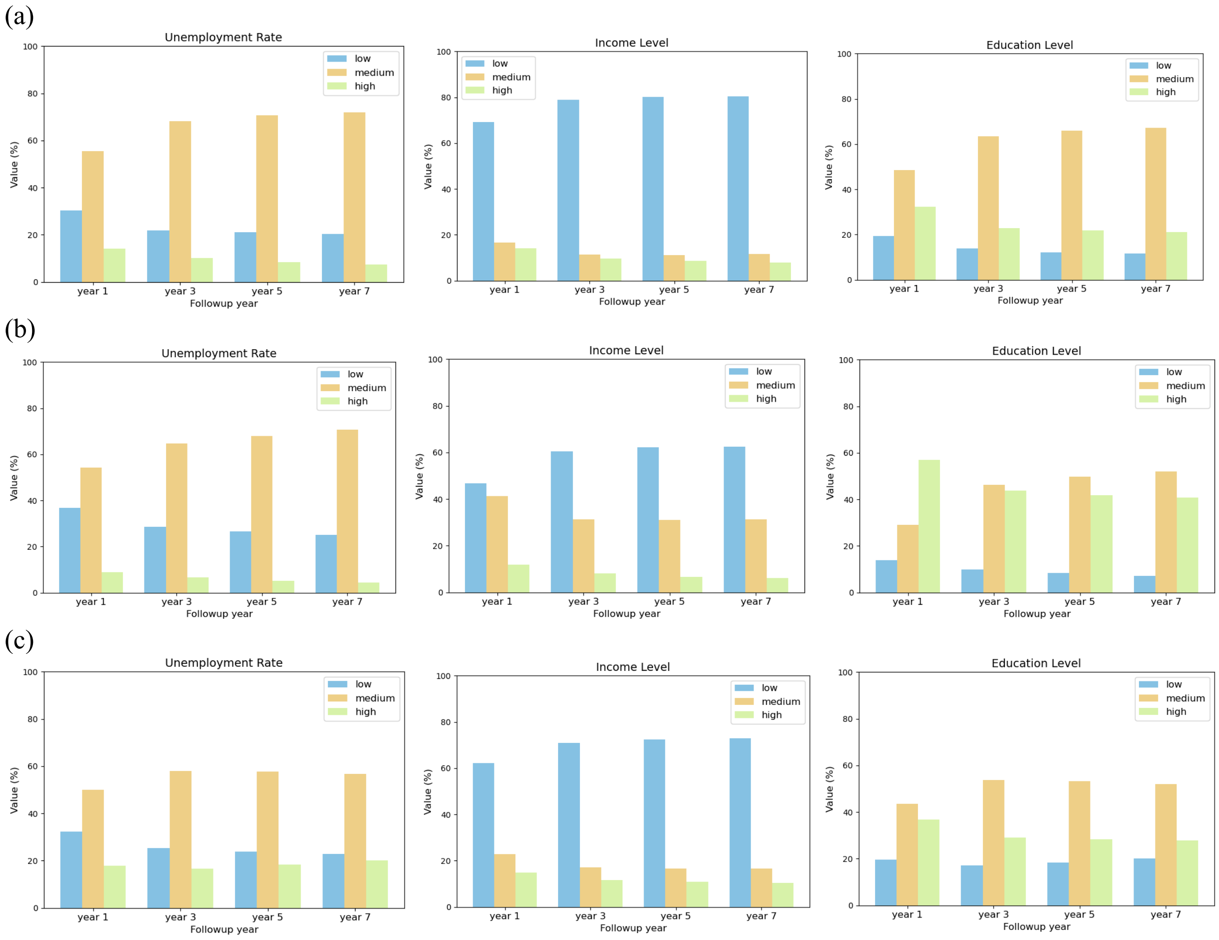


**eFigure 11. Opioids usage among subphenotypes.**

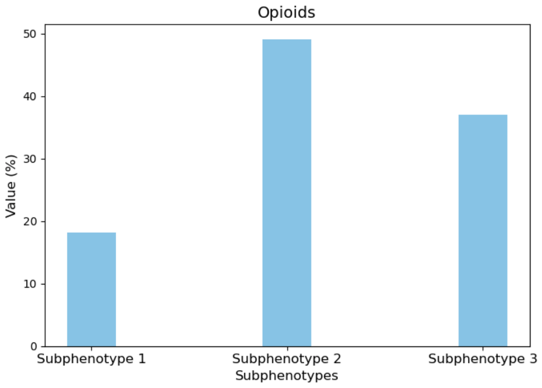
